# Automated quality control of T1-weighted brain MRI scans for clinical research: methods comparison and design of a quality prediction classifier

**DOI:** 10.1101/2024.04.12.24305603

**Authors:** Gaurav Bhalerao, Grace Gillis, Mohamed Dembele, Sana Suri, Klaus Ebmeier, Johannes Klein, Michele Hu, Clare Mackay, Ludovica Griffanti

## Abstract

**Introduction:** T1-weighted MRI is widely used in clinical neuroimaging for studying brain structure and its changes, including those related to neurodegenerative diseases, and as anatomical reference for analysing other modalities. Ensuring high-quality T1-weighted scans is vital as image quality affects reliability of outcome measures. However, visual inspection can be subjective and time-consuming, especially with large datasets. The effectiveness of automated quality control (QC) tools for clinical cohorts remains uncertain. In this study, we used T1w scans from elderly participants within ageing and clinical populations to test the accuracy of existing QC tools with respect to visual QC and to establish a new quality prediction framework for clinical research use.

**Methods:** Four datasets acquired from multiple scanners and sites were used (*N* = 2438, 11 sites, 39 scanner manufacturer models, 3 field strengths – 1.5T, 3T, 2.9T, patients and controls, average age 71 ± 8 years). All structural T1w scans were processed with two standard automated QC pipelines (MRIQC and CAT12). The agreement of the accept-reject ratings was compared between the automated pipelines and with visual QC. We then designed a quality prediction framework that combines the QC measures from the existing automated tools and is trained on clinical datasets. We tested the classifier performance using cross-validation on data from all sites together, also examining the performance across diagnostic groups. We then tested the generalisability of our approach when leaving one site out and explored how well our approach generalises to data from a different scanner manufacturer and/or field strength from those used for training.

**Results:** Our results show significant agreement between automated QC tools and visual QC (Kappa=0.30 with MRIQC predictions; Kappa=0.28 with CAT12’s rating) when considering the entire dataset, but the agreement was highly variable across datasets. Our proposed robust undersampling boost (RUS) classifier achieved 87.7% balanced accuracy on the test data combined from different sites (with 86.6% and 88.3% balanced accuracy on scans from patients and controls respectively). This classifier was also found to be generalisable on different combinations of training and test datasets (leave-one-site-out = 78.2% average balanced accuracy; exploratory models = 77.7% average balanced accuracy).

**Conclusion:** While existing QC tools may not be robustly applicable to datasets comprised of older adults who have a higher rate of atrophy, they produce quality metrics that can be leveraged to train a more robust quality control classifiers for ageing and clinical cohorts.

## Introduction

Large big brain MRI datasets hold immense value for well-powered statistical analyses and cross-cohort investigations (Madan, 2022). The emergence of open science initiatives and platforms for sharing data has made it possible to combine data from multiple sites and studies (Markiewicz et al., 2021; Wilkinson et al., 2016). With the emergence of comprehensive neuroimaging pipelines (e.g., UK Biobank, Human Connectome Project, etc.), it is now feasible to obtain imaging derived outcome measure on other datasets, including clinical populations (Littlejohns et al., 2020; Van Essen et al., 2013). In the ageing and dementia space there is a wealth of clinical datasets, made available through initiatives such as the Alzheimer’s Disease Neuroimaging Initiative (ADNI) and Dementias Platform UK (DPUK) (Bauermeister et al., 2020; Petersen et al., 2010). The aggregation of neuroimaging data obtained from clinical populations not only increases sample sizes but also facilitates the generation of reproducible and generalisable outcome measures, thus paving the way for innovative approaches in detecting brain biomarkers (Khanna et al., 2018; Van Horn & Toga, 2009). A substantial focus of neuroimaging research revolves around enhancing automated pipelines to produce reliable and relevant outcome measures from extensive datasets (Esteban et al., 2019; Frazier-Logue et al., 2022; Notter et al., 2023; Sherif et al., 2014). However, analysing large-scale datasets requires robust automated pipelines to ensure the generation of consistent measures across varied datasets. Despite the benefits, dealing with clinical datasets pose an additional challenge in big data analysis due to higher heterogeneity, motion artefacts, and disease-related factors like atrophy or other abnormalities (Andre et al., 2015; Nárai et al., 2022). Consequently, the critical task arises of identifying useable scans for processing through the automated pipelines to obtain reliable results.

While the MRI protocol may vary across datasets, a core component is a structural T1-weighted (T1w) scan. T1w MRI is used to examine brain structures, assess brain volume changes, and detect abnormalities, for example those associated with neurodegenerative diseases. It is also used as anatomical reference for the analysis of other structural and functional imaging modalities, as it provides detailed anatomical information. The initial and crucial step in brain imaging analysis involves assessing the quality of T1w MRI scans. The effectiveness of subsequent steps, such as multimodal registration and morphometry estimation, relies heavily on the quality of these scans. Traditionally, researchers visually inspect scans before analysis, but this practice isn’t always feasible when dealing with large datasets. Removing too many scans after quality assessment can decrease the sample size, while including poor-quality scans can introduce biases into the resulting outcomes (Gilmore et al., 2021).

Several automated approaches have been developed for quality control (QC) on T1w brain MRI scans (Hendriks et al., 2023). Various rule-based QC approaches have been proposed considering the image background to assess scan quality e.g. using measures such as - distortion (Woodard & Carley-Spencer, 2006), noise and ghosting artifacts (Gedamu et al., 2008), derived from image background (Mortamet et al., 2009), etc. Other rule-based QC approaches considered the image foreground to assess quality of the scans (Jang et al., 2018; Osadebey et al., 2018). Several automated machine learning approaches have been proposed, which extract quality measures from the images and are trained using visual QC labels to predict scan quality (pass or fail)(Alfaro-Almagro et al., 2018; Esteban et al., 2017; Pizarro et al., 2016). Various other studies used deep learning approaches to classify the scans as pass or fail using the entire image instead of specific quality measures (Bottani et al., 2022; Keshavan et al., 2019). Tools for brain morphometric analysis like Computational Anatomy Toolbox (CAT12) also offer quality control ratings based on tissue segmentation to evaluate scan quality (Gaser et al., 2022). While current automated QC tools are valuable, they are usually designed using data from healthy and/or young population or optimised for a specific dataset or type of scanner. To perform successful quality control in large clinical datasets, it is important to establish a framework that offers broader applicability across various clinical cohorts, age range and scanner types.

In this study, we tested two existing automated QC tools: MRIQC and CAT12. MRIQC is an open-source tool, offering an extensive array of metrics for evaluating quality on raw T1w images (based on noise, information theory, and specific artifacts), and it has become a standard reference in numerous studies (Chen et al., 2023; Elliott et al., 2023; Lorenzini et al., 2022). CAT12 is widely utilized in the field and encompasses a variety of quality control options (based on noise contrast, inhomogeneity contrast, resolution) applicable to images processed within the tissue segmentation pipeline (Besteher et al., 2022; Hahn et al., 2022; Sakreida et al., 2022). To classify the scans into pass or fail, MRIQC additionally provides a pre-trained supervised classifier which can be utilised to predict the quality of scans. In contrast CAT12 provides image quality ratings for each measure which can be used to determine usable or unusable scans from the analysis. Due to their wide use and broad range of comprehensive measures available in both tools from raw and tissue-segmented scans, we selected these tools as good candidates to perform QC on clinical datasets. We first tested the agreement between MRIQC and CAT12 with visual quality inspection on a large sample of clinical research data (*N* = 2438) from an extensive spectrum of datasets spanning ageing and neurodegenerative cohorts. We studied the relationship between the QC metrics produced by the two tools and tested the tools’ performance when adjusting the accept-reject threshold. We then proposed a new classification framework which uses a combination of QC metrics from both automated tools as features and visual QC as gold standard. We tested the generalisability of the proposed classifier on various test datasets that differed in terms of population and scanner. Finally, by looking at the distribution of QC measures that contributed most to the higher classification accuracy, we explored how they could be used to inform data harmonisation strategies. The code is openly available, and the proposed classifier will be made accessible on the DPUK data portal, to support future clinical research studies.

## Methods

### Data & visual QC of T1w brain scans

Structural T1w brain images from 4 clinical research datasets (*N* = 2438) acquired on 39 scanners from three different manufacturers (Siemens, Philips, GE) were used: 1) Oxford Brain Health Clinic (BHC) (Griffanti et al., 2022) [age range: 65 - 101 years], 2) Oxford Parkinson’s Disease Centre (OPDC) (Griffanti et al., 2020) [age range: 39 - 116 years], 3) Whitehall II imaging study (Filippini et al., 2014) [age range: 60 – 85 years], 4) Alzheimer’s Disease Neuroimaging Initiative (ADNI) (Petersen et al., 2010) [age range: 55 - 92 years]. Information on scanner, manufacturing model, counts, acquisition matrix and voxel size for these datasets is provided in **Table 1**.

**Table 1.**
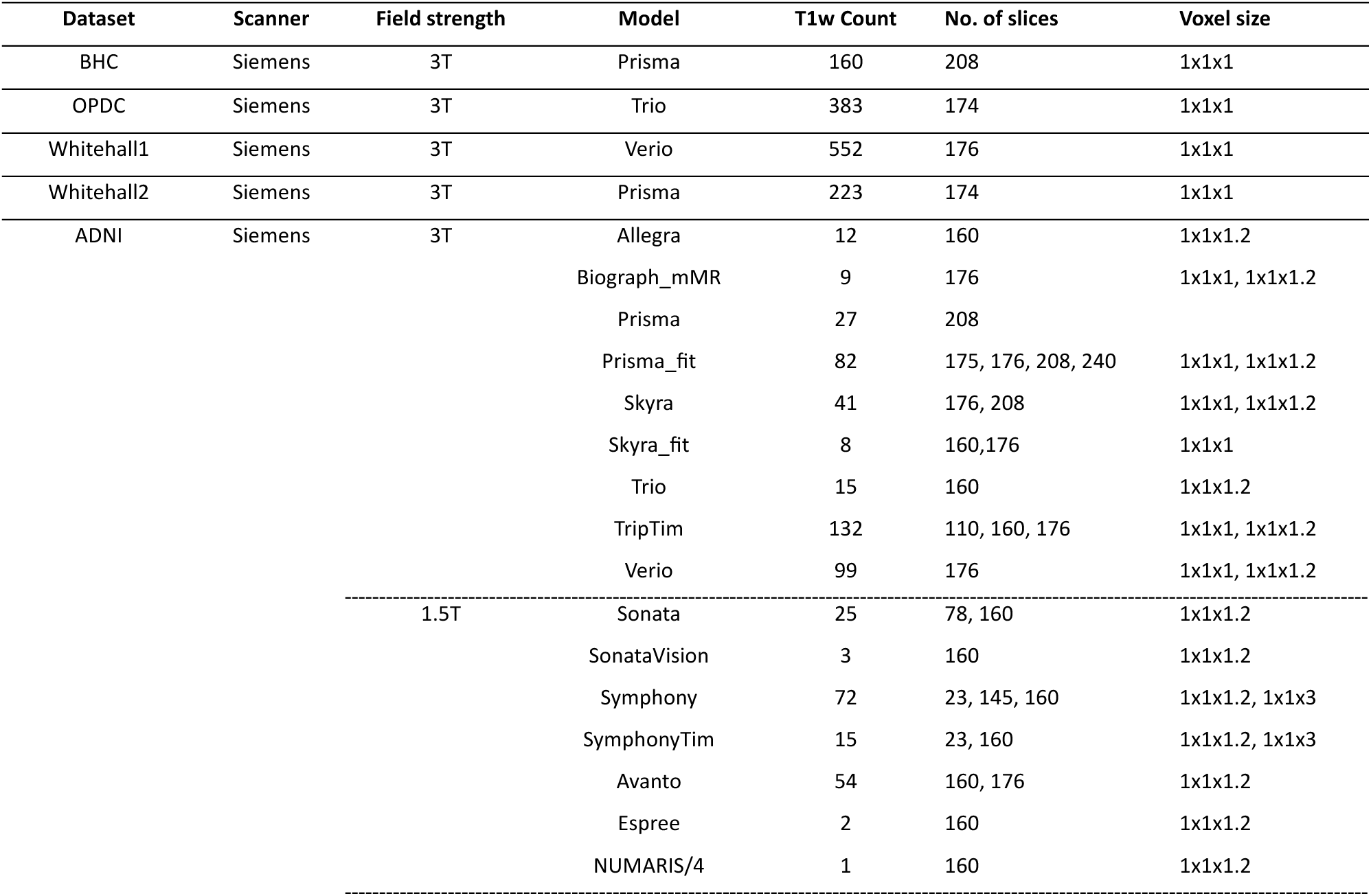
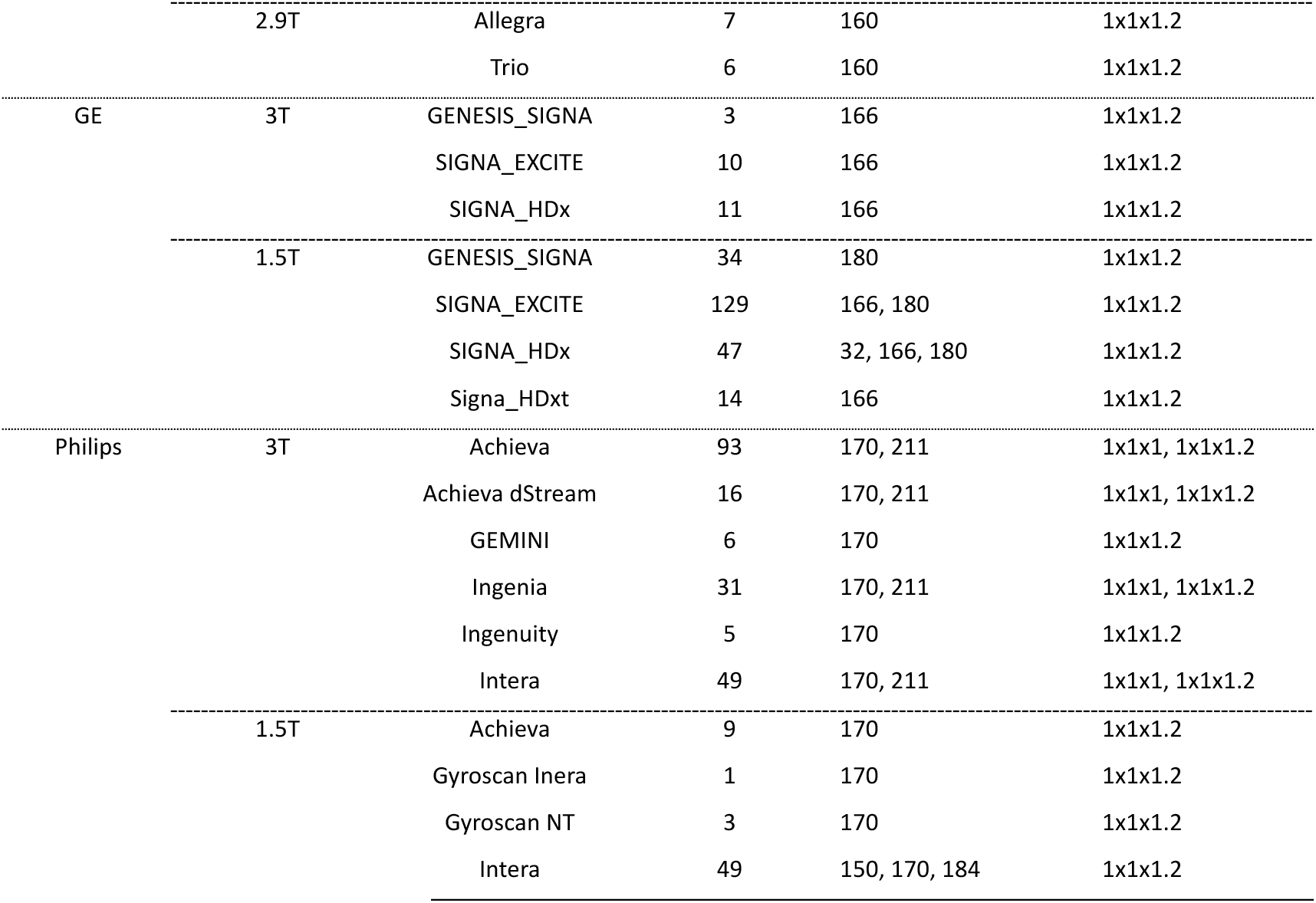
Dataset-wise and scanner-wise counts of T1w scans of datasets used in this study.

### Oxford Brain Health Clinic - BHC *(N=160)*

The Oxford BHC is a joint clinical-research service for memory clinic patients which offers high-quality assessments not routinely available, including a multimodal brain MRI scan (Griffanti et al., 2022). Images are acquired on a Siemens 3T Prisma scanner using a protocol matched with the UK Biobank imaging study (Miller et al., 2016). The visual quality ratings were obtained from the dataset owners. These images were originally rated into low, medium, high quality. We categorised medium and high-quality images into accept label and low-quality images into reject label.

### Oxford Parkinson’s Disease Centre Discovery Cohort - OPDC *(N=383)*

The OPDC study aims to identify biomarkers of Parkinson’s disease for early detection and progression. The dataset includes multimodal brain MRI data (acquired on a 3T Siemens Verio scanner) along with deep longitudinal clinical phenotyping in patients with Parkinson’s, at-risk individuals, and healthy elderly volunteers (Griffanti et al., 2020). For this dataset, the visual ratings were not available from dataset owners hence each image was visualised and rated into low, medium, and high quality by one rater. The medium and high-quality images were grouped into accept category and low-quality images were in reject category.

### Whitehall II imaging sub-study (*N=775*)

The Whitehall II study is a longitudinal study of British civil servants to explore the factors affecting brain health and cognitive ageing (Filippini et al., 2014). In this dataset, 552 scans were acquired on a Siemens Verio 3T scanner (referred as Whitehall1 in the manuscript – protocol details in ((Filippini et al., 2014) and 223 scans on a Siemens Prisma 3T (referred as Whitehall2 in the manuscript – protocol details in (Zsoldos et al., 2020)). We treated the data from these two scanners separately in all the analyses for our work. The visual quality ratings (accept and reject) were obtained from the dataset owners.

#### Alzheimer’s Disease Neuroimaging Initiative - ADNI *(N=1120)*

The ADNI (adni.loni.usc.edu) was launched in 2003 as a public-private partnership, led by Principal Investigator Michael W. Weiner, MD. The primary goal of ADNI has been to test whether serial MRI, positron emission tomography (PET), other biological markers, and clinical and neuropsychological assessment can be combined to measure the progression of mild cognitive impairment (MCI) and early Alzheimer’s disease (AD). In this study we included all the baseline T1w brain images from ADNI 1,2,3 and GO (first run in each session). Due to the highly variable numbers of scans for each scanner, we grouped data from the same manufacturer and field strength together, for a total of 7 ADNI sites. The visual quality ratings were available on a scale from *1* (excellent quality) to *4* (unusable). Upon careful inspection of the quality description, we decided to label images with a rating of 1 or 2 into the accept category and those with a rating of 3 or 4 into the reject category.

### T1w processing in automated tools

All the images were named and organised in Brain Imaging Data Structure (BIDS) (Gorgolewski et al., 2016) and defaced (to preserve the privacy of individuals) before processing.

### MRIQC pipeline

MRIQC is an open-source pipeline that extracts image quality metrics (IQMs) from structural (T1w and T2w) and functional MRI data (Esteban et al., 2017). It uses modular sub-workflows from neuroimaging software toolboxes such as *FSL* (Jenkinson et al., 2012), *ANTs* (Avants BB et al., 2013)and *AFNI* at the background (Cox & Hyde, 1997). MRIQC also provides a random forest classifier (mriqc_clf) pre-trained on 1102 T1w scans (17 sites) from the Autism Brain Imaging Data Exchange (ABIDE) dataset. The classifier generates probability value for each scan (range *0* - *1*) and any scan with probability more than or equal to 0.5 (default threshold) is categorised to reject label.

Each defaced T1w brain image was processed in MRIQC pipeline (singularity version 0.15.1). The list of image quality metrics (IQMs) and their description are provided in **Table 2** (a detailed explanation can be found in the user manual of MRIQC). From each image 68 metrics were extracted. We used MRIQC’s random forest classifier (mriqc_clf) and labelled images into binary accept and reject labels.

**Table 2:**
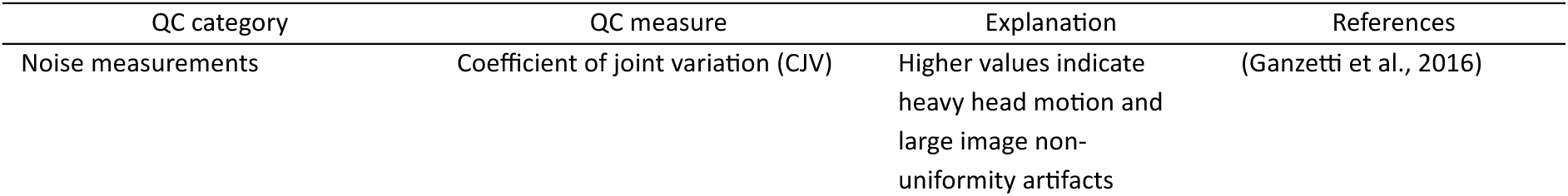
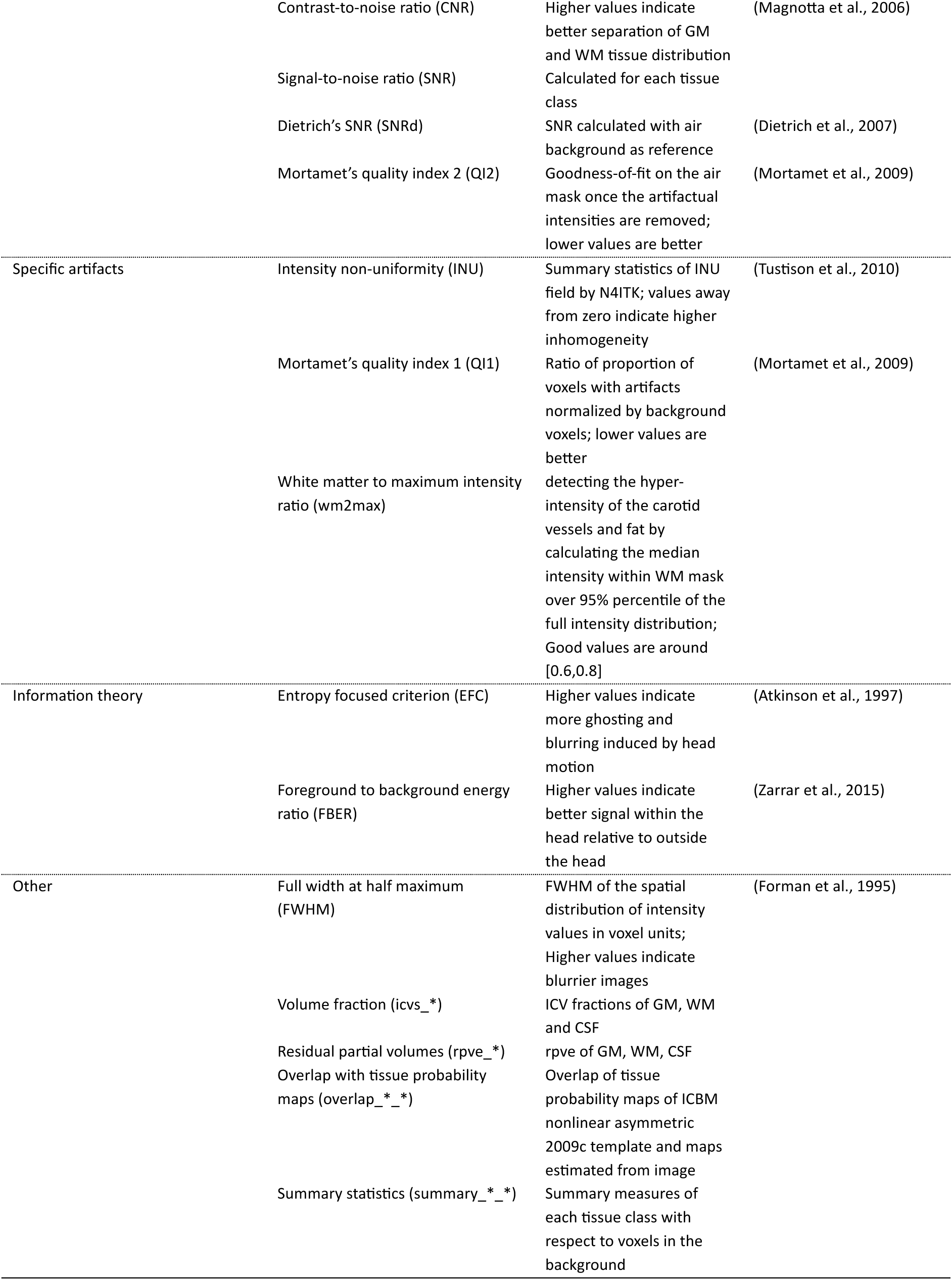
List of MRIQC image quality metrics.

### CAT12 pipeline

CAT12 (Computational Anatomy Toolbox) is an extension of SPM12 covering diverse morphometric methods to provide computational anatomy (Gaser et al., 2022). CAT12 provides a retrospective QC framework for empirical quantification of image quality parameters.

Each defaced T1w brain image was processed in CAT12 segmentation pipeline (standalone version r2042 running on v93 of MATLAB compiler runtime). The surface processing option was enabled during the segmentation. Post segmentation, CAT12 generates a segmentation report for each image and provides image quality ratings (IQRs) based on noise, resolution, bias and aggregates these ratings to weighted IQR [range *A+* (excellent) to *F* (unacceptable/failed)]. Additionally, (for the proposed classifier work) we also considered additional quality measures which are not provided in the CAT12 visualisation report but saved in the output of segmentation (named as, *cat_<subjdirname>.mat*). The description of all the quality measures is provided in **Table 3**, (a detailed explanation can be found in the user manual of CAT12). From each image 36 quality measures were extracted (Pravesh Parekh, 2021). To label the images into accept and reject quality, each image with weighted image quality rating (IQR) of C minus and below (selected as ‘default threshold’) was labelled into reject class.

**Table 3.**
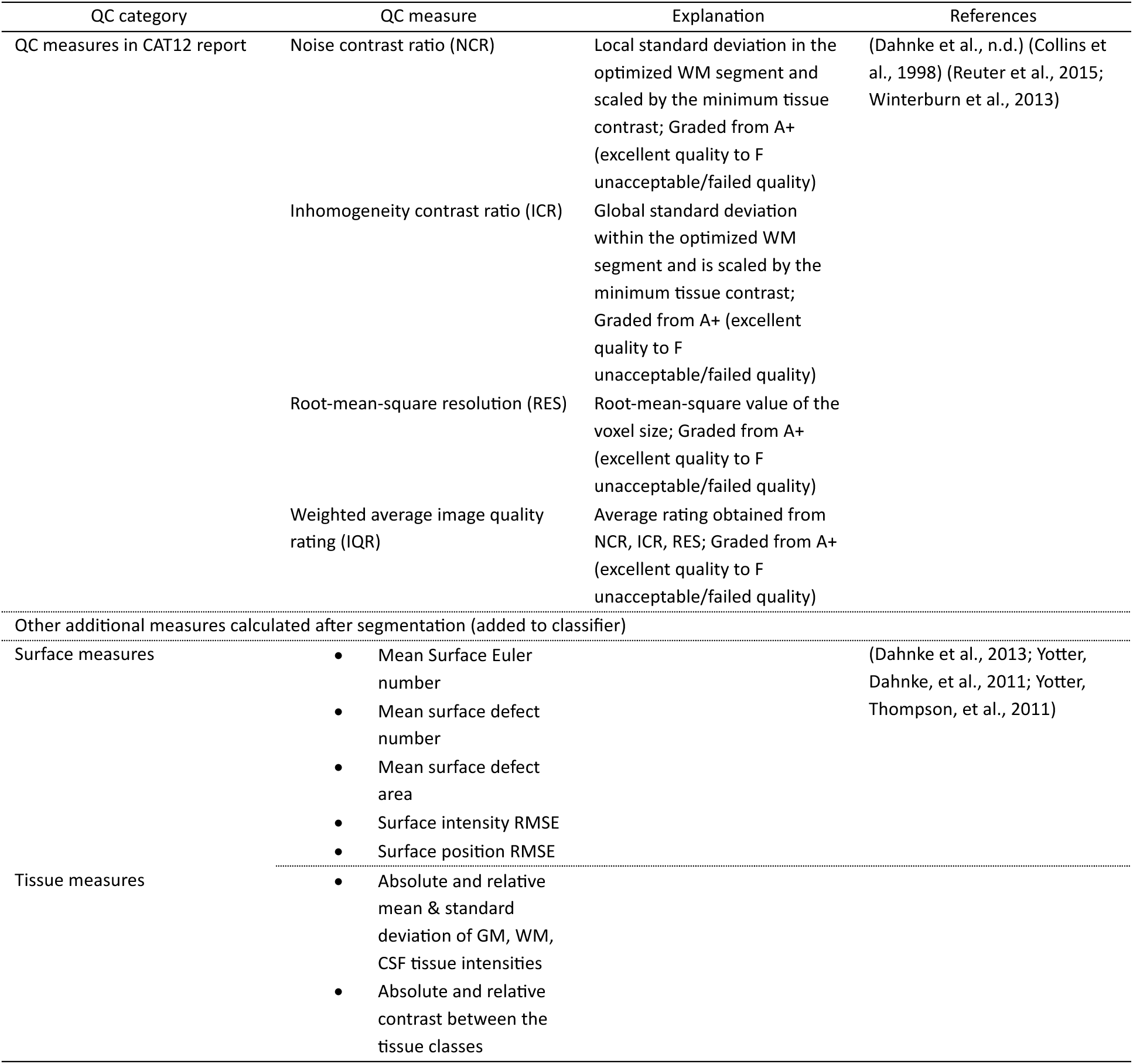
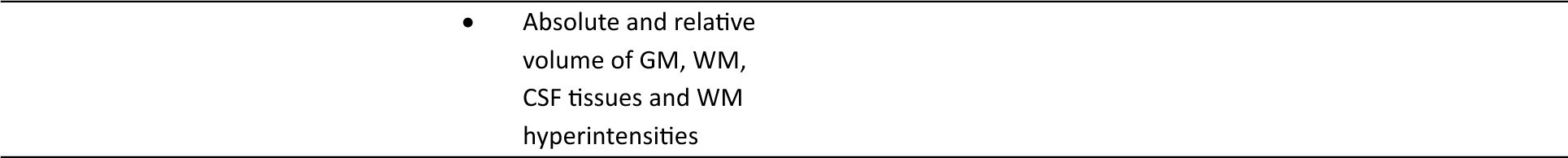
List of CAT12 image quality measures.

### Comparison of MRIQC and CAT12 quality measures

We first compared the quality measures between the two automated tools. The quality measures derived from MRIQC and CAT12 were correlated using Pearson’s correlation. The correlation analysis was conducted in MATLAB2022b (*The MathWorks Inc. (2022). MATLAB Version: 9.13.0.2105380 (R2022b)*, 2022).

### Comparison of ratings between automated tools and visual QC

We calculated the percentage of scans that would pass QC and compared the agreement between visual QC, MRIQC classifier predictions (default threshold, *MRIQC(D)*) and CAT12’s weighted IQR (default threshold, *CAT12(D)*) using Kappa coefficient of inter-rater reliability (IRR) (Landis & Koch, 1977; McHugh, 2012). Three comparisons were performed: 1) CAT12 ratings vs. MRIQC ratings, 2) CAT12 ratings vs. visual QC ratings, 3) MRIQC ratings vs. visual QC ratings.

Further, we explored the effect of changing the labelling threshold from MRIQC classifier and CAT12’s weighted IQR. We investigated this by changing the CAT12’s weighted IQR threshold to – 1) strict (*CAT12 (-)*): any scan with weighted IQR rating C and below were labelled to reject category, 2) lenient (*CAT12 (+)*): any scan with weighted IQR rating D+ and below were labelled to reject category. Similarly, for the MRIQC classifier we changed the threshold of acceptance to – 1) strict (*MRIQC (-)*): scans with probability equal to or more than 0.4 were labelled to reject category, 2) lenient (*MRIQC (+)*): scans with probability equal to or more than 0.6 were labelled to reject category. We then re-calculated the Kappa coefficient for the above three comparisons. The Kappa coefficient was calculated using IRR package in R (Matthias Gamer et al., 2019; R Core Team, 2022).

### Proposed QC classifier

In this section we present our proposed QC classifier. The primary model (combined data model) was trained and tested on a mix of data from multiple datasets and sites. We then tested the generalisability of our classification framework in a leave-one-site-out approach and in cases where training and test data differ in terms of field strength and/or scanner manufacturer.

### Combined data model

#### Data and classifiers

We designed a binary QC classifier which combines the MRIQC and CAT12 quality measures as features. Binary visual QC ratings were used as target. For the combined data model, we first randomly divided our entire sample (*N* = 2438) into 80% training (*N* = 1955) and 20% test data (*N* = 483). The data was divided ensuring fair representation of target labels, sites, and proportion of patients and controls (when applicable) among both the training and test datasets. The site-wise and label-wise split for training and test datasets is provided in **Table 4**. We tested three options for the underlying machine learning classification: support vector machine, random forest, and random under-sampling boost.

**Table 4.** Site-wise split of training and test data for the combined data model.

| Datasets | Train data |  |  | Test data |  |  |
| --- | --- | --- | --- | --- | --- | --- |
|  | Reject | Accept | Total | Reject | Accept | Total |
| ADNI GE 1.5T | 8 | 172 | 180 | 1 | 43 | 44 |
| ADNI GE 3T | 4 | 16 | 20 | 0 | 4 | 4 |
| ADNI Philips 1.5T | 1 | 49 | 50 | 0 | 12 | 12 |
| ADNI Philips 3T | 7 | 153 | 160 | 2 | 38 | 40 |
| ADNI Siemens 3T | 8 | 332 | 340 | 2 | 83 | 85 |
| ADNI Siemens 1.5T | 10 | 128 | 138 | 2 | 32 | 34 |
| ADNI Siemens 2.9T | 2 | 9 | 11 | 0 | 2 | 2 |
| OPDC Siemens 3T | 47 | 260 | 307 | 12 | 64 | 76 |
| BHC Siemens 3T | 12 | 116 | 128 | 4 | 28 | 32 |
| Whitehall1 Siemens 3T | 35 | 407 | 442 | 9 | 101 | 110 |
| Whitehall2 Siemens 3T | 6 | 173 | 179 | 1 | 43 | 44 |
| Total | 140 | 1815 | 1955 | 33 | 450 | 483 |

Support vector machine (SVM) is one of the most common supervised classifiers, simple to train for hyperparameters, effectively handles high dimensional data and less prone to overfitting than non-linear classifiers (Cortes & Vapnik, 1995). We used the ‘fitcsvm’ implementation in MATLAB (*MATLAB Version 9.14.0.2239454 (R2023a)*, 2023). Two hyperparameters were optimised in nested cross-validation (CV): box constraint (0.01, 0.1, 1, 10, 100, 1000) and Kernel function (linear, radial basis function). The remaining hyperparameters were maintained at their default settings. Random forest (RF) is a supervised classifier robust to outliers and non-linear data, faster to train and handles unbalanced classes in the data (as in our data ‘reject’ class samples are substantially lower than ‘accept’ class) (Breiman, 2001). We used the ‘fitcensemble’ implementation in MATLAB (*MATLAB Version 9.14.0.2239454 (R2023a)*, 2023). Two hyperparameters were optimised in nested CV: Maximal number of decision splits (10,50) and number of ensemble learning cycles (10, 50, 100). The remaining hyperparameters were maintained at their default settings. We selected random under-sampling boost (RUS) as third classifier due to its ease of implementation, effective handling of imbalanced classes, rapid processing speed, and reduced computational complexity (Seiffert et al., 2008). It is a supervised classifier that under samples the majority class labels in the training process to balance the minority class. Given the imbalance of classes in our data, we used random under-sampling to avoid skewing towards the majority class (accept) and improve the detection of the minority class (reject) in our datasets. We used the ‘fitcensemble’ implementation in MATLAB (*MATLAB Version 9.14.0.2239454 (R2023a)*, 2023). Three hyperparameters were optimized in nested CV: Maximal number of decision splits (10, 50), number of ensemble learning cycles (10, 50, 100), and learning rate for shrinkage (0.01, 0.1). The remaining hyperparameters were maintained at their default settings.

#### Nested cross validation approach

The classifiers were trained in a nested cross validation (CV) framework consisting 5 outer folds and 3 inner folds (See **Figure 1**). In the training phase, within every CV iteration, the features were standardized for each site separately. During the feature standardization of test data, only the mean and standard deviation from train data were applied to avoid data leakage. Within the CV, features were ranked using multiple filter-type feature selection methods (ReliefF, Chi-square, Minimum Redundancy Maximum Relevance, class separability criteria – t-test, entropy, Bhattacharya distance, Wilcoxon, Receiver Operating Characteristics). The ranks were then aggregated using robust ranking aggregation (Kolde et al., 2012). For each feature size (iterative; 10, 20, 30, 40, 50, 60, 70, 80, 90, 100, 104 (all)), the classifier was trained on the inner fold’s train data and tested on the inner fold’s test data for the grid of hyperparameters. For each outer CV iteration and for each feature size, the classification performance was averaged over all the inner CV folds and the combination of hyperparameters achieving the best performance were chosen. Finally, for each feature size the outer cross validation iteration was executed with the chosen combination of hyperparameters from the inner folds, models were re-trained, and tested on the outer test data. To get precise estimates of model’s performance, we ran a total 100 iterations of the nested CV in the training phase and obtained the best combination of hyperparameters for each feature size for each classifier. In the final model design, we aggregated feature ranks from all outer cross validation folds across 100 iterations and derived a final ranking of the features. The final model was then trained by using all the training data with the best combination of hyperparameters for each feature size and feature ranking across 100 CV iterations.

**Figure 1.**
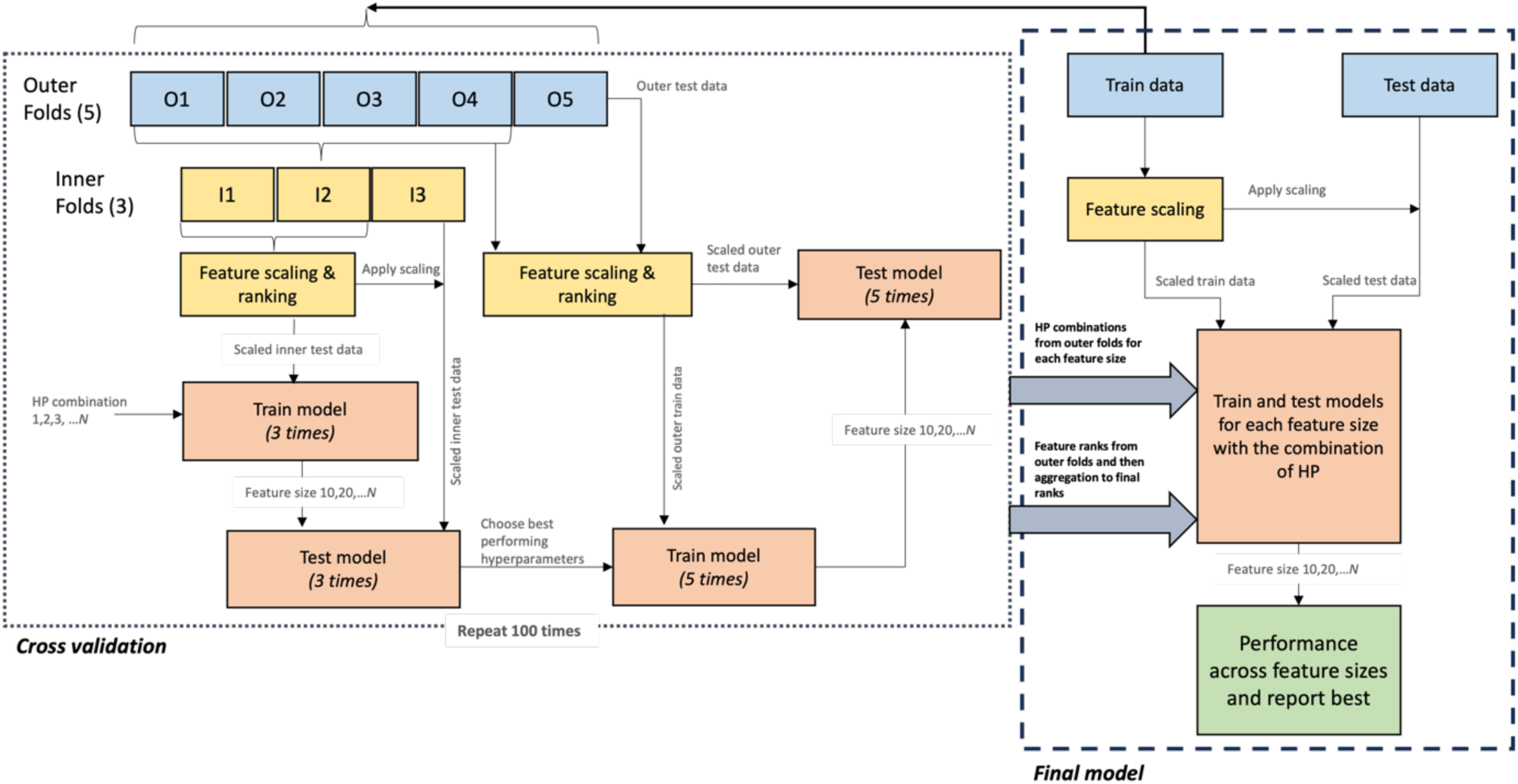
Nested cross validation workflow for training the QC classifier. The model was trained for 5 outer folds and 3 inner folds. The hyperparameters of the model were optimised on the inner test data and the combination giving the best performance (balanced accuracy) were selected for the outer folds. The nested cross validation process was repeated for 100 times. The best performing hyperparameters for each feature size and feature ranks across 100 iterations were used to train the final model and tested on the hold-out data.

#### Assessment and comparison of prediction performance

The final model’s performance on the test data was assessed by balanced accuracy (**Eq. 1 – 3**).

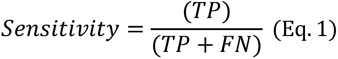

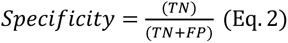

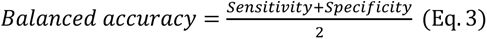

*True positive (TP) – the model correctly predicts the accept label; True negative (TN): correctly predicts reject label; False positive (FP) – wrongly predicts accept label for a scan that should be rejected; False negative (FN) – wrongly predicts reject label a scan that should be accepted*.

The choice to use balanced accuracy as our primary metric is based on the fact that our datasets have imbalance in the accept and reject classes and we are interested in both classes being predicted well for unseen datasets.

For each classifier (SVM, RF, RUS) we selected the feature size that gave the best performance. We then compared the prediction performance of the three optimised classifiers with each other and with MRIQC and CAT12. This comparison of prediction performance was done for - 1) combined test data (*N* = 483), 2) test data categorised by site (see **Table 4** for number of scans in each site in each class), 3) patients and controls separately within the test data (see **Table 5** for number of scans for in each class), 4) each sub-category of diagnosis within the test data (see **Table 5** for number of scans in each class for each category).

**Table 5.** Diagnosis group-wise number of scans in accept and reject labels.

|  | Diagnosis group | Reject | Accept | Total |
| --- | --- | --- | --- | --- |
| Controls | ADNI | 2 | 71 | 73 |
|  | OPDC | 2 | 14 | 16 |
|  | Whitehall1 | 9 | 101 | 110 |
|  | Whitehall2 | 1 | 43 | 44 |
|  | Total | 14 | 229 | 243 |
| Patients | ADNI Dementia | 2 | 26 | 28 |
|  | ADNI MCI | 3 | 117 | 120 |
|  | OPDC RBD | 4 | 23 | 27 |
|  | OPDC iPD | 6 | 27 | 33 |
|  | BHC | 4 | 28 | 32 |
|  | Total | 19 | 221 | 240 |

*MCI: mild cognitive impairment, RBD: REM sleep behaviour disorder (at risk group for PD), iPD: idiopathic Parkinson's disease*

#### Feature importance

We investigated the distribution of the top 10 ranked features derived from the final combined data model by employing kernel density and scatter plots. To ascertain potential statistical variations in the distribution among sites, we conducted a two-sample Kolmogorov-Smirnov test. Subsequently, to address multiple comparisons, we applied the Bonferroni correction to obtain adjusted p-values.

### Leave-one-site out models

To further validate our approach, we created leave-one-site out CV models using the best performing classifier on the combined data (among SVM, RF and RUS) to see how well our training workflow generalises to an unseen site. From this classifier, we extracted the combinations of hyperparameters, feature ranking and feature size at which the best performance was observed on the combined test data. These parameters were then used to re-train classifier on data from remaining sites while keeping each site as test data. Finally, the classification performance on each test site was assessed, comparing them against MRIQC and CAT12, and against the best performance of a combined data model on each site in the test data. The split of data for training and testing for each model is provided in **Table 6**.

**Table 6.** Train and test split for leave-one-site-out models.

| Test Dataset | Train Reject | Train Accept | Train Total | Test Reject | Test Accept | Test Total |
| --- | --- | --- | --- | --- | --- | --- |
| ADNI GE 1.5T | 132 | 1643 | 1775 | 9 | 215 | 224 |
| ADNI GE 3T | 136 | 1799 | 1935 | 4 | 20 | 24 |
| ADNI Philips 1.5T | 139 | 1766 | 1905 | 1 | 61 | 62 |
| ADNI Philips 3T | 133 | 1662 | 1795 | 9 | 191 | 200 |
| ADNI Siemens 3T | 132 | 1483 | 1615 | 10 | 415 | 425 |
| ADNI Siemens 1.5T | 130 | 1687 | 1817 | 12 | 160 | 172 |
| ADNI Siemens 2.9T | 138 | 1806 | 1944 | 2 | 11 | 13 |
| OPDC Siemens 3T | 93 | 1555 | 1648 | 59 | 324 | 383 |
| BHC Siemens 3T | 128 | 1699 | 1827 | 16 | 144 | 160 |
| Whitehall1 Siemens 3T | 105 | 1408 | 1513 | 44 | 508 | 552 |
| Whitehall2 Siemens 3T | 134 | 1642 | 1776 | 7 | 216 | 223 |

### Exploratory models

Finally, we explored how well our approach generalises when the model is trained on data from one field strength and/or manufacturer and tested on data from other field strengths/manufacturers. These exploratory models were designed to test:

1. Generalisability across field strength: the majority of the datasets were acquired on 3T scanners (*N* = 1967) hence we trained the model on data from all 3T scanners and tested on the data from 1.5T field strengths.
2. Generalisability across manufacturer: the majority of the datasets were acquired from Siemens scanners (*N*=1928) hence we trained the model combining Siemens data from all field strengths and tested on data from other manufacturers.
3. Generalisability across manufacturer and field strength: the majority of the data is from 3T Siemens scanners (*N* = 1743) hence we trained the model only from 3T Siemens scanner data and tested on the remaining data.

The data split for training and test data is provided in **Table 7**. Similar to the leave-one-site out models, we chose the best performing classifier (among SVM, RF and RUS) on the combined test data and re-trained and tested the classifier for three different cases. As explained above, the training process used hyperparameter combinations, feature ranking and feature size at which the best performance was observed on the combined test data. The classification performances were assessed for each model, comparing them against MRIQC and CAT12, and against the performance of the combined data model on the test data from each case individually.

**Table 7.** Training and test data split for exploratory models.

| Models | Training sites | *N* training – Total<br>(accept) | Test dataset | *N* test – Total<br>(accept) |
| --- | --- | --- | --- | --- |
| Generalisability<br>across field<br>strength | 3T (Siemens,<br>Philips, GE) | 1576 (1457) | Siemens 1.5T,<br>Philips 1.5T, GE<br>1.5T | 458 (436) |
| Generalisability<br>across<br>manufacturer | 3T, 2.9T, 1.5T<br>(Siemens) | 1545 (1425) | 3T (Philips, GE),<br>1.5T (Philips, GE) | 510 (487) |
| Generalisability<br>across<br>manufacturer and<br>field strength | 3T (Siemens) | 1396 (1288) | 3T (Philips, GE),<br>1.5T (Siemens,<br>Philips, GE), 2.9T<br>(Siemens) | 695 (658) |

## Results

### Comparison of quality measures (CAT12 vs MRIQC)

We analysed the correlation between CAT12 quality measures and MRIQC IQMs (**Figure 2**) to explore both common and distinct metrics within these tools. We observed various statistically significant correlation coefficients between pairs of measures from these automated tools. For instance, CAT12’s resolution measure exhibited significant correlation with MRIQC’s summary-based metrics derived from tissues, FWHM, image size, image spacing, and overlap of CSF with tissue probability maps (TPM). The absolute volume of tissues measured by CAT12 demonstrated significant correlations with MRIQC’s intra-cranial volume fraction of tissues and overlap of tissue classes with TPM. The relative intensity of background in CAT12 exhibited a significant correlation with MRIQC measures encompassing noise-based metrics, measures tied to specific artifacts (such as image nonuniformity), as well as other parameters like size, spacing, FWHM, and residual partial volumes of tissues. Relative intensities from CSF and GM in CAT12 were significantly correlated with summary measures from background, CSF, and GM in MRIQC. Summary measures derived from WM in MRIQC significantly correlated with CAT12’s resolution, relative intensity of CSF, and absolute volumes of GM and WM tissues. Similarly, CAT12’s relative contrast showed a significant correlation with summary measures from CSF and GM in MRIQC. On the other hand, non-significant or low correlations (below ±0.5) suggest that the two tools are also capturing unique information about the image. Measures falling in this category for CAT12 are noise, mean intensity from tissues, and surface measures while for MRIQC are QI1, QI2 (targeting specific artifacts), EFC, FBER (informed by information theory). For detailed correlation coefficients, p-values, and the upper and lower bounds of a 95% confidence interval for each pair of measures, refer to Supplementary material.

**Figure 2.**
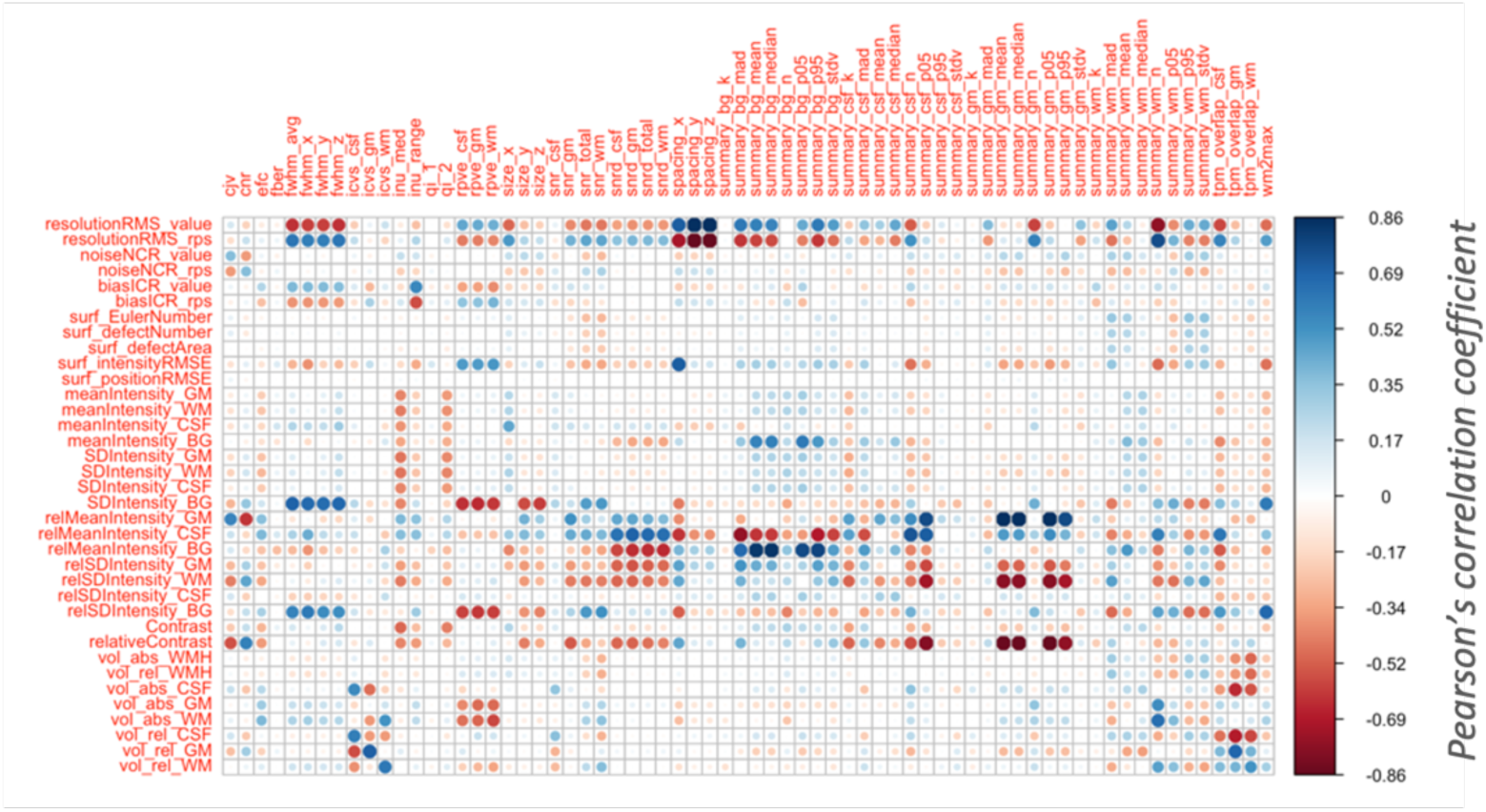
Correlation plot between MRIQC IQMS (columns) and CAT12 quality measures (rows). MRIQC generated total 68 IQMs and from CAT12 we extracted 36 quality measures.

### Comparison of ratings between automated tools and visual QC

#### Percentage of scans passing QC

We first compared the percentage of scans that passed (accept category) QC using visual QC, MRIQC, and CAT12. The results are reported in **Table 8**. Overall, CAT12 showed the highest percentage of accepted scans compared to visual QC and MRIQC. MRIQC showed a more similar percentage of accepted scans to visual QC overall, but with over 8% difference in 3 datasets (ADNI 3T GE, ADNI 1.5T Philips and BHC)

**Table 8.** Percentage of scans passing visual QC and QC from automated tools.

| Dataset | Field Strength | Scanner | Total N scans available | % accept Visual QC | % accept MRIQC (MRIQC – Visual) | % accept CAT12 QC (CAT12 – Visual) |
| --- | --- | --- | --- | --- | --- | --- |
| ADNI | 1.5T | GE | 224 | 96 | 96 (0) | 98.7 (2.7) |
| ADNI | 3T | GE | 24 | 83.3 | 91.7 (8.4) | 95.8 (4.1) |
| ADNI | 1.5T | Philips | 62 | 98.4 | 90.3 (-8.1) | 100 (9.7) |
| ADNI | 3T | Philips | 200 | 95.5 | 88 (-7.5) | 98 (10) |
| ADNI | 3T | Siemens | 425 | 97.6 | 94.1 (-3.5) | 98.8 (4.7) |
| ADNI | 1.5T | Siemens | 172 | 93 | 89 (-7.7) | 98.8 (9.8) |
| ADNI | 2.9T | Siemens | 13 | 84.6 | 76.9 (-7.7) | 100 (23.1) |
| OPDC | 3T | Siemens | 383 | 84.6 | 91.4 (6.8) | 98.7 (7.3) |
| BHC | 3T | Siemens | 160 | 90 | 80 (-10) | 91.9 (11.9) |
| Whitehall1 | 3T | Siemens | 552 | 92 | 92.9 (0.9) | 94.6 (1.7) |
| Whitehall2 | 3T | Siemens | 223 | 96.9 | 96.4 (-0.5) | 98.7 (2.3) |
| All datasets |  |  | 2438 | 92.9 | 91.8 | 97.3 |

#### Classification agreement

We computed Kappa coefficient to measure the agreement between the automated tools and with visual QC (**Figure 3**). A detailed table of Kappa coefficients, associated p-values, and percentage agreement for all the pairs of ratings is provided in supplementary material.

**Figure 3.**
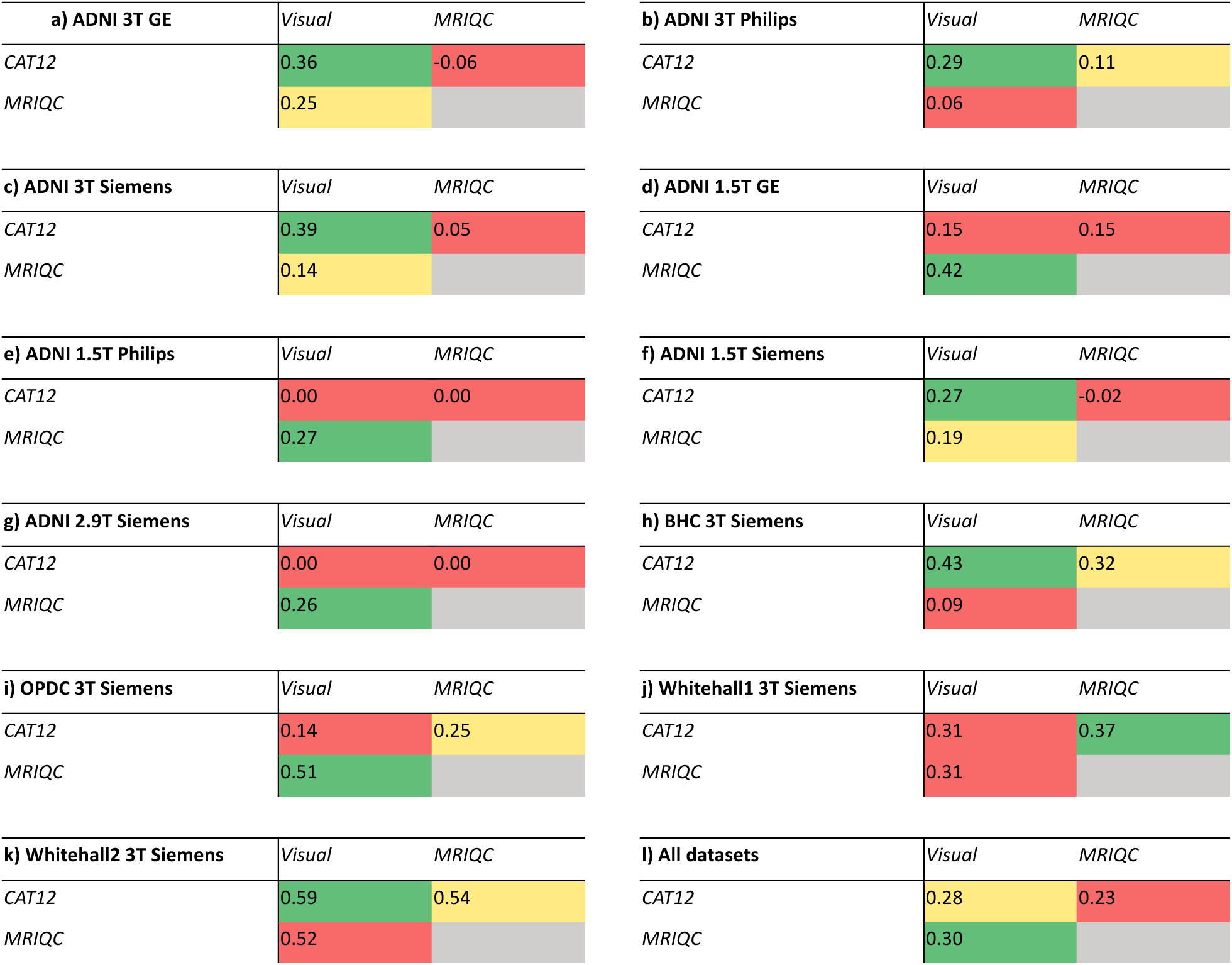
Kappa coefficient values comparing the agreement of ratings between visual QC and automated tools. The colours indicate the lowest (red), medium (yellow) and highest values (green) in each dataset. See supplementary material for details.

#### Automated tools vs visual QC

When evaluating the agreement on all datasets together (**Figure 3** panel l), MRIQC and visual QC showed higher value of Kappa coefficient (k=0.3) than CAT12 and visual QC (k=0.28). However, when looking at each dataset separately, in some cases the agreement was higher between CAT12 and visual QC (panels a, b, c, f, h, k, Kappa between 0.27 and 0.59), while other datasets showed higher Kappa coefficient between MRIQC and visual QC (panels d, e, g, i, j, Kappa between 0.26 and 0.51). Notably, ADNI 1.5T Philips and 2.9T Siemens showed no agreement between CAT12 and visual QC (panels e, g).

#### CAT12 vs MRIQC ratings

For all datasets together, we found significant agreement between the ratings from CAT12 and MRIQC ratings (k=0.23). When considered each dataset separately, Whitehall2 dataset showed the highest agreement (k=0.54). Notably, some datasets in ADNI (3T GE, 1.5T Siemens, 1.5T Philips, 2.9T Siemens) showed no agreement or worse than expected agreement (zero or negative values of Kappa coefficient in Figure 4 panels a, f, e, g).

**Figure 4.**
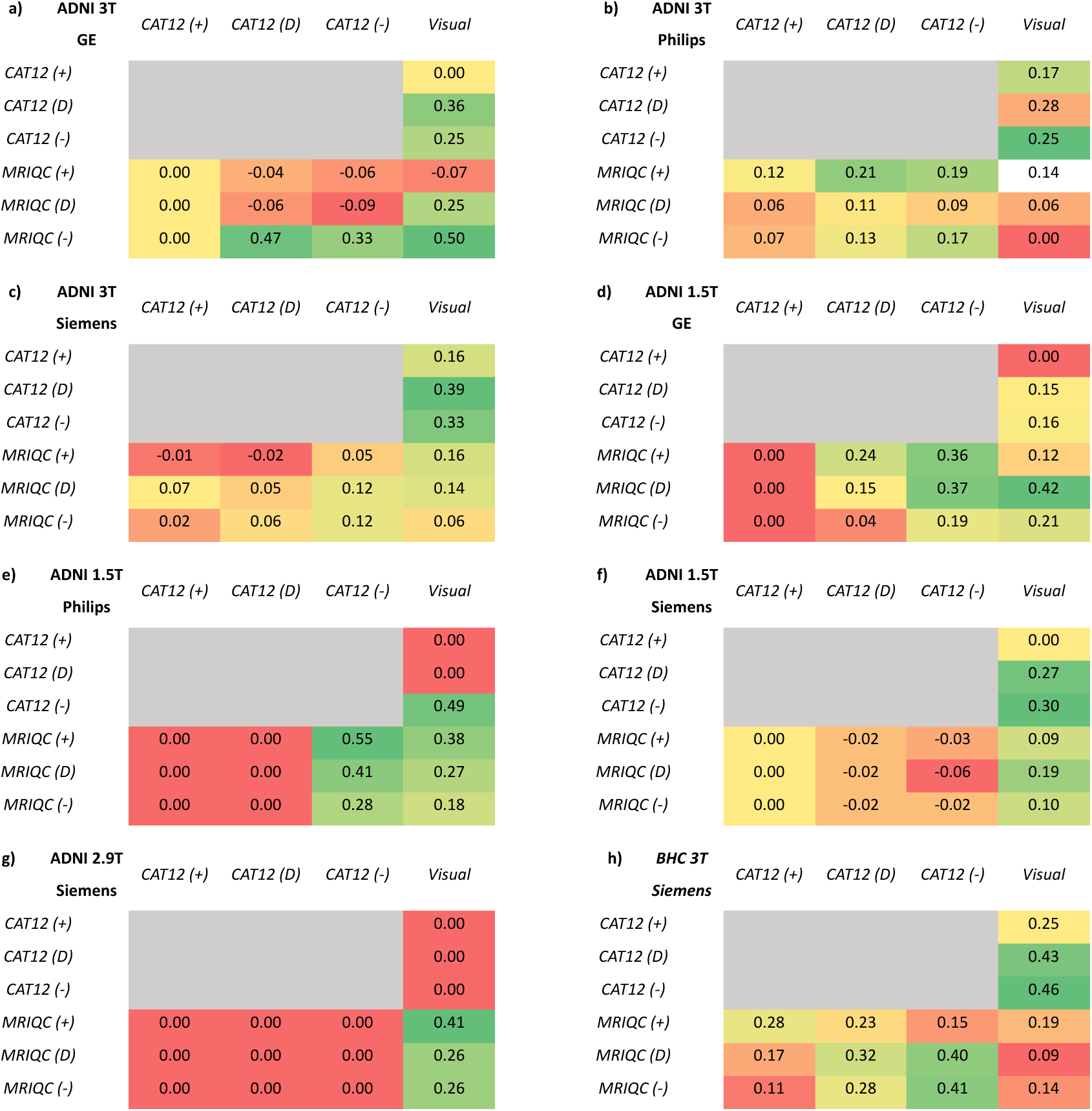
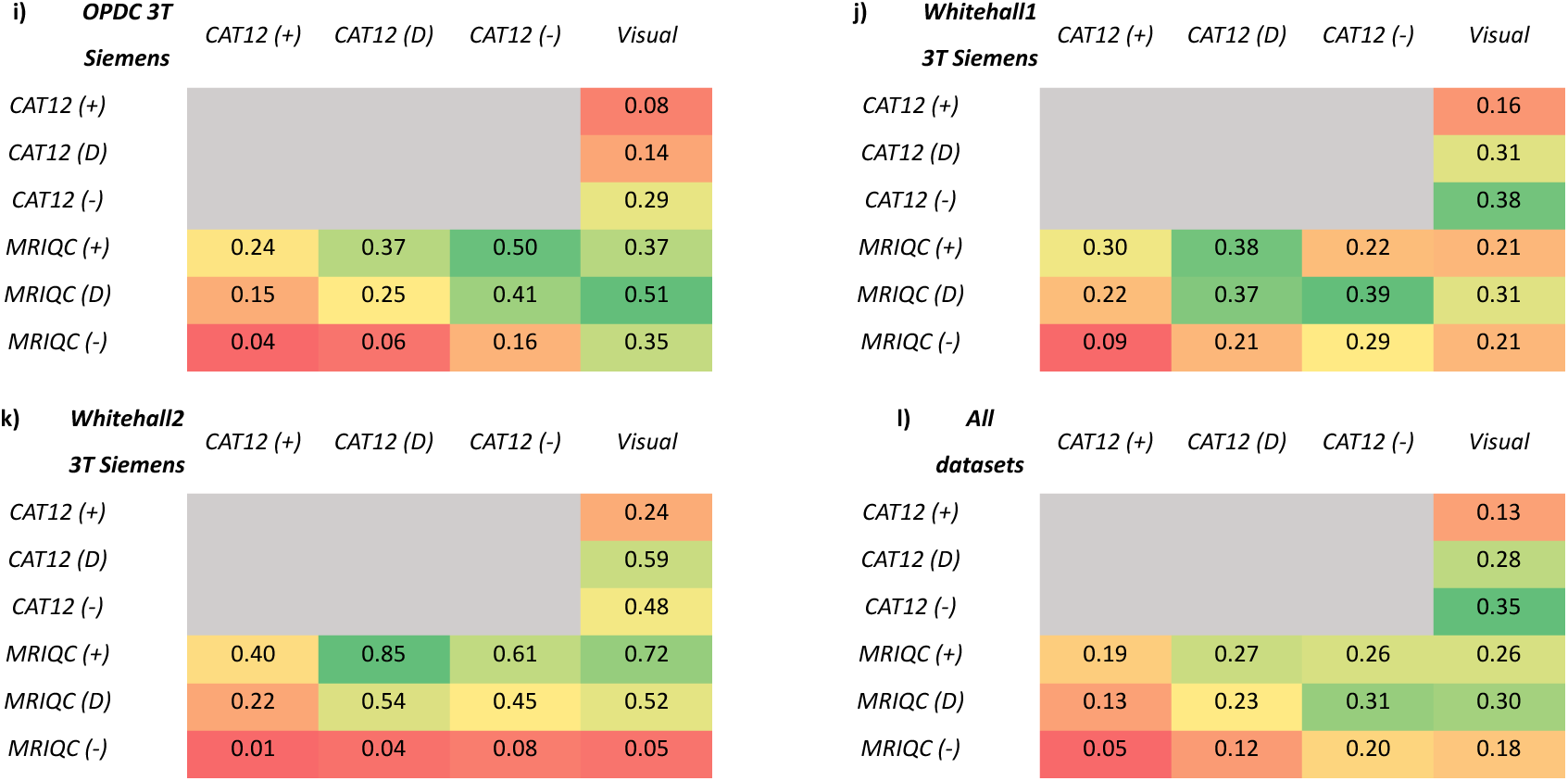
Kappa coefficient values comparing the agreement of ratings between visual QC and automated tools after adjusting the acceptance thresholds of automated tools. The comparison with default thresholds is also provided for ease of comparison. The colours indicate the lowest (red), medium (yellow) and highest values (green) in each panel. See supplementary material for details.

#### Impact of threshold on classification agreement

Given that the inter-rater reliability did not show consistency across datasets on which tool produced more similar ratings to visual QC using their default threshold (0.28 for CAT12, 0.30 for MRIQC), we explored the effect of using a lenient and stricter threshold of acceptance on the automated tools. The percentage of accepted scans upon adjusting the threshold are provided in **Table 9**. A detailed table of Kappa coefficients, associated p-values, and percentage agreement for all the pairs of ratings is provided in Supplementary material.

**Table 9.** Percentage of accepted scans after adjusting acceptance thresholds for MRIQC and CAT12 (‘-‘ for strict threshold and ‘+’ for lenient threshold).

| Dataset | Field Strength | Scanner | Total scans | % accept Visual QC | % accept CAT12 (-) (CAT12 (-) – Visual QC) | % accept CAT12 (+) (CAT12 (+) – Visual QC) | % accept MRIQC (-) (MRIQC (-) – Visual QC) | % accept MRIQC (+) (MRIQC (+) – Visual QC) |
| --- | --- | --- | --- | --- | --- | --- | --- | --- |
| ADNI | 1.5T | GE | 224 | 96 | 95.1 (1.1) | 100 (4) | 86.6 (-10.6) | 97.8 (1.8) |
| ADNI | 3T | GE | 24 | 83.3 | 91.7 (8.4) | 100 (16.7) | 87.5 (4.2) | 95.8 (12.5) |
| ADNI | 1.5T | Philips | 62 | 98.4 | 95.2 (-3.2) | 100 (1.6) | 85.5 (-12.9) | 93.5 (-4.9) |
| ADNI | 3T | Philips | 200 | 95.5 | 94 (-1.5) | 99 (3.5) | 78 (-17.5) | 93.5 (-2) |
| ADNI | 3T | Siemens | 425 | 97.6 | 96.9 (-0.7) | 99.5 (1.9) | 83.5 (-14.1) | 97.2 (-0.4) |
| ADNI | 1.5T | Siemens | 172 | 93 | 96.5 (3.5) | 100 (7) | 77.3 (-15.7) | 97.7 (4.7) |
| ADNI | 2.9T | Siemens | 13 | 84.6 | 100 (15.4) | 100 (15.4) | 76.9 (-7.7) | 84.6 (0) |
| OPDC | 3T | Siemens | 383 | 84.6 | 95.6 (11) | 99.2 (14.6) | 69.7 (-14.9) | 94.5 (9.9) |
| BHC | 3T | Siemens | 160 | 90 | 79.4 (-10.6) | 96.9 (6.9) | 65 (-25) | 91.3 (1.3) |
| Whitehall1 | 3T | Siemens | 552 | 92 | 88.2 (-3.8) | 98.2 (6.2) | 75.7 (-16.3) | 97.3 (5.3) |
| Whitehall2 | 3T | Siemens | 223 | 96.9 | 96 (-0.9) | 99.6 (2.7) | 61.4 (-35.5) | 98.2 (1.3) |
| All datasets |  |  | 2438 | 92.9 | 93 (0.1) | 99 (6) | 75.8 (-17.1) | 96.1 (3.2) |

#### Visual QC vs automated tools

We recalculated the Kappa coefficient values after adjustment of thresholds to find the agreement between the automated tools and visual QC ratings (see **Figure 4**). When looking at the Kappa coefficient values from all datasets together (panel l), we found that the agreement between visual QC and CAT12 ratings improved after applying a strict threshold to CAT12. We found a similar effect when looking at each dataset separately on most of our datasets (panels b, d, e, h, I, j). For example, the lack of agreement between visual QC ratings and default threshold ratings of CAT12 (k =0) for ADNI 1.5T Philips dataset improved significantly (k=0.43) after applying strict threshold to CAT12 ratings. However, some datasets did not show any improvement in Kappa coefficient after adjusting thresholds (panels a, c, g, k).

For all datasets together, we did not see any improvement in Kappa coefficient when comparing visual QC with changed thresholds in MRIQC ratings. Some datasets showed increased agreement after applying lenient threshold to MRIQC ratings (panels b, c, e, g, h, k). For example, the significant agreement between visual QC and default threshold ratings of MRIQC in Whitehall2 (k = 0.52) was further improved (k = 0.72) after applying lenient threshold to MRIQC ratings. Only ADNI 1.5T GE dataset showed significantly improved value of Kappa coefficient after applying strict threshold to MRIQC ratings (from k = 0.25 to k= 0.5). The rest of the datasets did not show any improvement upon adjusting the threshold of MRIQC ratings (panels d, f, i, j).

#### CAT12 vs MRIQC ratings

For all datasets together and each dataset separately, the Kappa coefficient significantly improved between MRIQC default threshold ratings and CAT12 ratings after applying a strict threshold (from k=0.23 to k=0.31). Most of the datasets showed similar effect of improvement between default threshold ratings of MRIQC and CAT12 ratings after applying strict threshold (panels c, d, e, h, i, j). For Whitehall2 and ADNI 3T Philips, the Kappa coefficient improved between default ratings of CAT12 and MRIQC ratings after applying lenient threshold. Only for ADNI 3T GE, the Kappa coefficient value between default threshold ratings of CAT12 and MRIQC (from k =-0.06 to 0.47) after applying strict threshold to MRIQC ratings. Notably, ADNI 1.5T Siemens and ADNI 2.9T Siemens did not show any improvement upon adjustment of thresholds, showing zero agreement.

### Classification performance

#### Combined data model

The optimal feature size selected for SVM (balanced accuracy = 67.4%) and RF was 50 (balanced accuracy =72.5%), while for RUS was 80 (balanced accuracy = 87.7%) (**Figure 5**). On an average across different feature sizes for the combined test data, the proposed RUS classifier showed the highest balanced accuracy (85.2 ± 2.8%) as compared to SVM (62.8 ± 4.9%) and RF classifier (65.8 ± 3.7%) (refer to supplementary material for details for performance at each feature size and confusion matrices for each classifier and automated tools). The comparison of the best performance of the proposed classifiers with MRIQC and CAT12 showed that CAT12 (56.9%) gave the lowest balanced accuracy on the test data as compared to all classifiers while MRIQC (71.6%) showed higher balanced accuracy than SVM but lower than RF and RUS classifiers.

**Figure 5.**
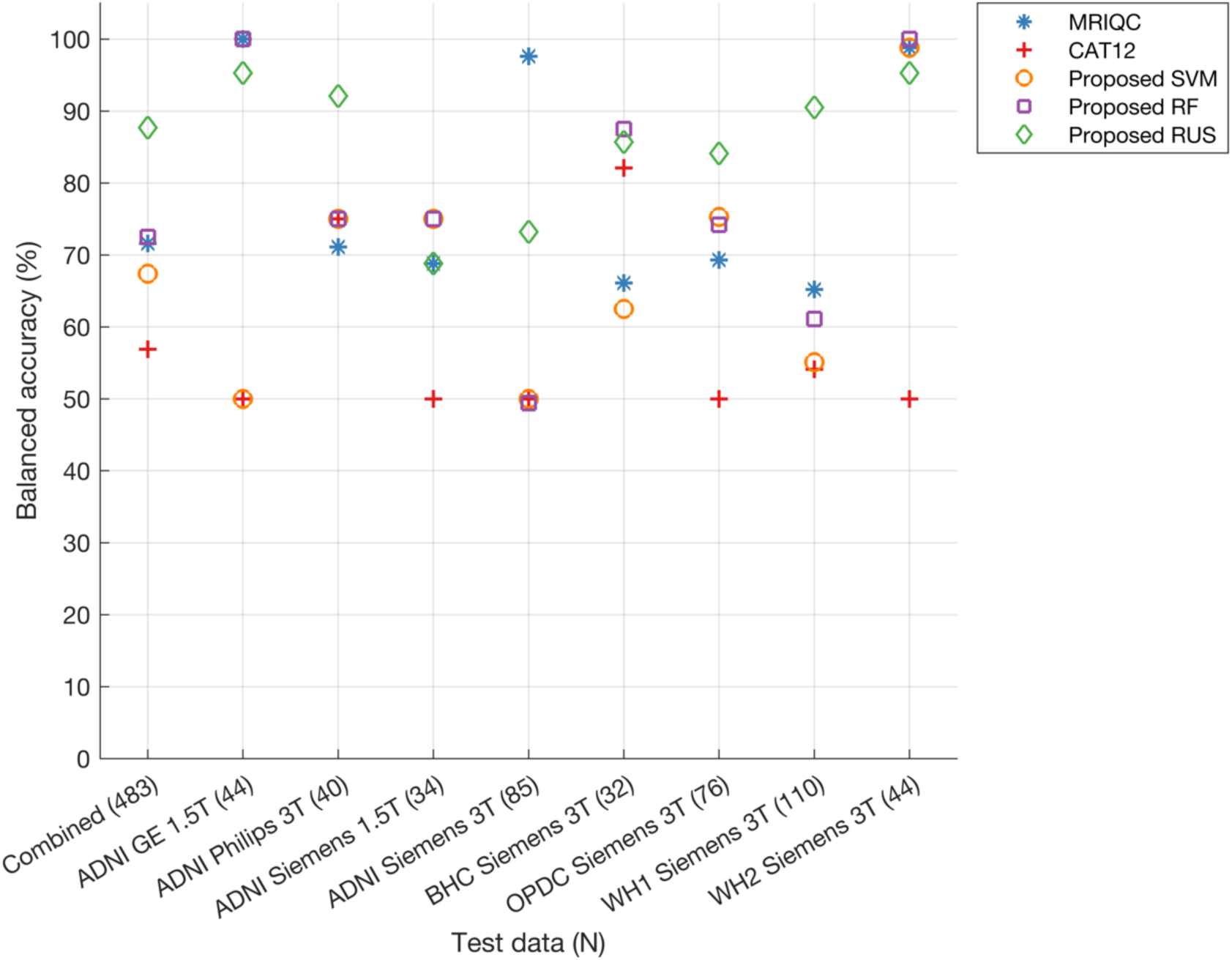
Balanced accuracy of proposed classifiers, MRIQC and CAT12 on combined and site-wise test data. Number of samples in the test data are provided in brackets for each dataset (x-axis). Note that three sites (ADNI GE 3T, ADNI Philips 1.5T and ADNI Siemens 2.9T) are not included in the figure due to the absence of samples in the reject class resulting in NaN values for balanced accuracies.

When looking at the performance for each site separately in the test data (**Figure 5**), the proposed classifiers showed higher balanced accuracies compared to CAT12 (except for BHC Siemens 3T where CAT12 showed higher balanced accuracy only when compared to SVM). We found that RUS achieved the highest balanced accuracies for 3 sites (ADNI Philips 3T, OPDC Siemens 3T and Whitehall1 Siemens 3T sites). For other sites (ADNI GE 1.5T, ADNI Siemens 1.5T, BHC Siemens 3T, Whitehall2 Siemens 3T), either MRIQC or RF showed the highest balanced accuracies, but RUS performance was also very close. For ADNI Siemens 3T site, MRIQC showed the highest balanced accuracy (97.6%), followed by RUS classifier (73.2%).

#### Performance for patients and controls in test data

Since our aim is to have a classifier that is suitable for clinical data, we evaluated the performance separately for different disease groups. When grouping scans in broad categories of patients (N=240 in the test set, including AD, MCI, PD, RBD) and controls (N=243 in the test set, generally cognitively unimpaired and without neurological conditions), the RUS classifier (balanced accuracy: patients = 86.8%, controls = 88.3%) achieved superior performance as compared to both SVM (balanced accuracy: patients = 75.6%, controls = 56.3%) and RF classifier (balanced accuracy: patients = 78.7%, controls = 64.1%) and existing tools MRIQC (balanced accuracy: patients = 72.5%, controls = 69.9%) and CAT12 (balanced accuracy: patients = 59.8%, controls = 52.9%) (**Figure 6**).

**Figure 6.**
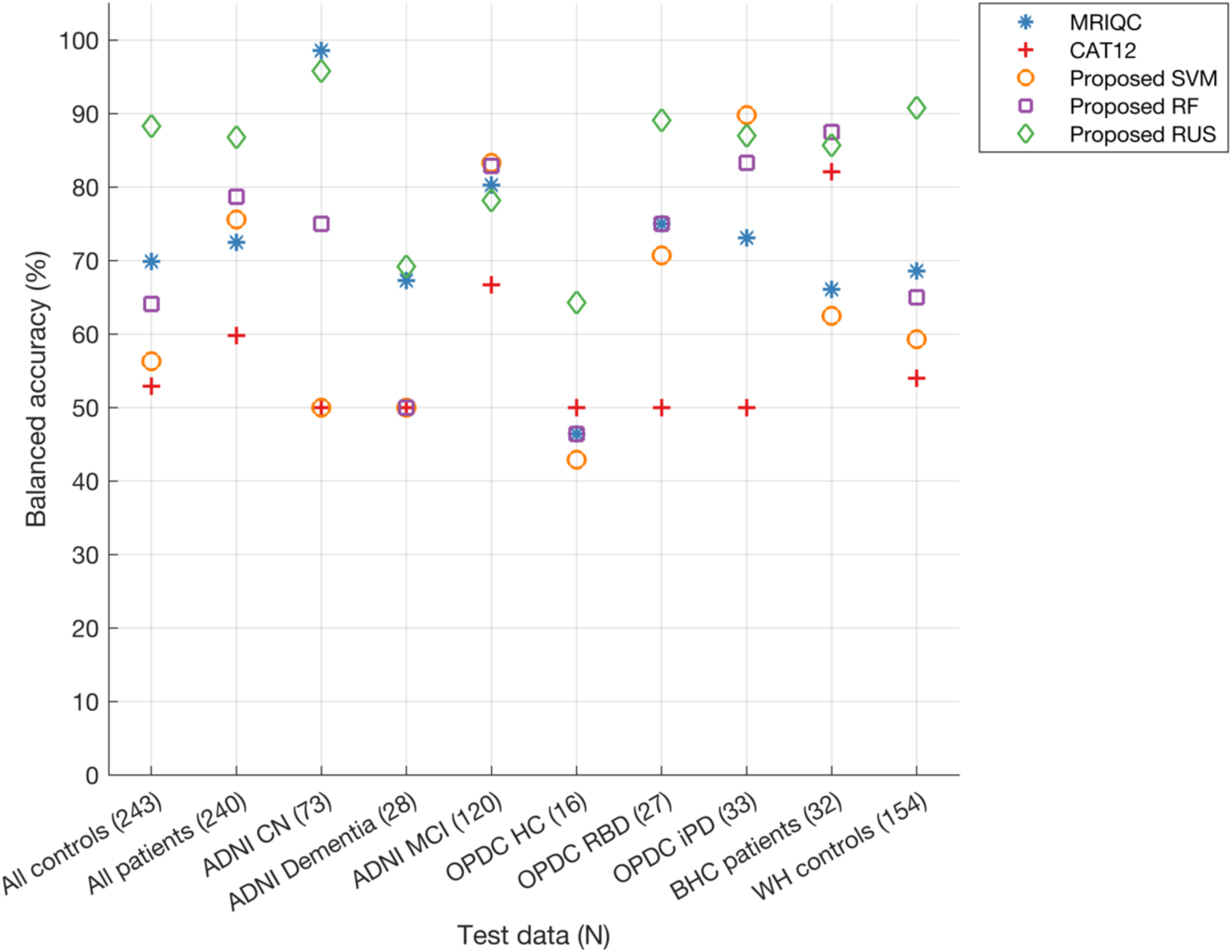
Balanced accuracy of proposed classifiers, MRIQC, and CAT12 analysed separately for scans from healthy individuals, patients, and each diagnostic sub-category within both healthy and patient groups in the test dataset. Number of samples in the test data are provided in brackets for each category (x-axis). Legend of diagnostic subgroups: CN = cognitively normal; HC = Healthy Controls; MCI = Mild Cognitive Impairment; RBD = REM Sleep Behaviour Disorder; iPD = idiopatic Parkinson’s Disease.

When looking at the performance for different diagnostic groups within ADNI, for the MCI group MRIQC, RF and SVM classifiers showed the highest balanced accuracy (>80%) while RUS accuracy was also very close to these classifiers (78%). The RUS classifier showed the highest balanced accuracy on the dementia group (69.2%) with MRIQC achieving 67.3%. On BHC data (memory clinic patients) the proposed SVM (87.5%) showed the highest balanced accuracy with the RUS classifier achieving 85.7%.

When looking at the performance for different diagnostic groups within OPDC, for the RBD group, the RUS classifier showed the highest balanced accuracy (89.1%). For the iPD group, the proposed SVM and RUS classifiers showed the highest balanced accuracies (>85%). Upon comparing balanced accuracies across control groups from various datasets, the proposed RUS classifier demonstrated superiority for most datasets, achieving 64.3% for OPDC HC and 90.8% for Whitehall II controls. The only exception is the ADNI CN group, where the MRIQC classifier achieved the highest balanced accuracy at 98.6% with the RUS classifier performing close to MRIQC (95.8%).

#### Feature importance

The feature ranking of the final model (combined data model) included features from both CAT12 and MRIQC in the top ranked features. The top 80 features (feature size showing the best balanced accuracy for the proposed RUS classifier) included 23 features from CAT12 [noise, contrast ratio, surface and tissue measures] and 57 features from MRIQC [summary measures, noise measures and tissue measures]. We selected the top 10 features (from 80 features) and plotted them to explore the distribution of these QC measures for each site in datasets (See KS density plots in **Figure 7** for different sites in ADNI and **Figure 8** for other sites). The plots reveal significant variations in the distribution of the top 10 features among different sites, highlighting technical variability despite the datasets originating from scanners of the same manufacturer. For instance, the disparity in feature distribution between the BHC dataset and others, despite all being acquired on 3T Siemens scanners, is evident (refer to **Figure 8** panels b, c, d, f, i, j). Conversely, the distribution of features in the ADNI dataset suggests a more consistent pattern across various sites (refer to **Figure 7** panels a to l).

**Figure 7.**
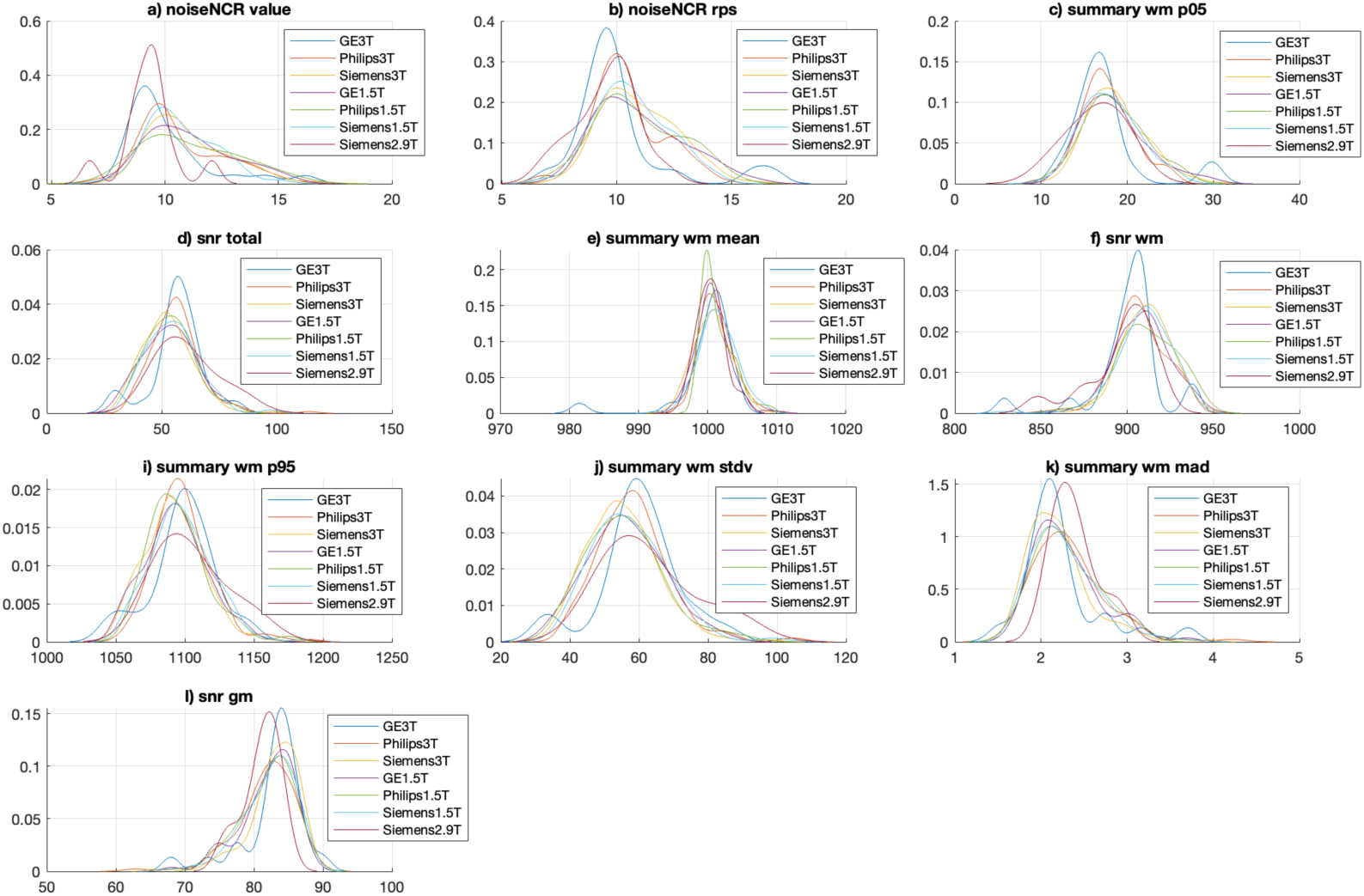
Kernel density plots showing the distribution of top 10 ranked features in final combined data model for sites within ADNI dataset. For a description of the features, please refer to tables **Table 2** and **Table 3**.

**Figure 8.**
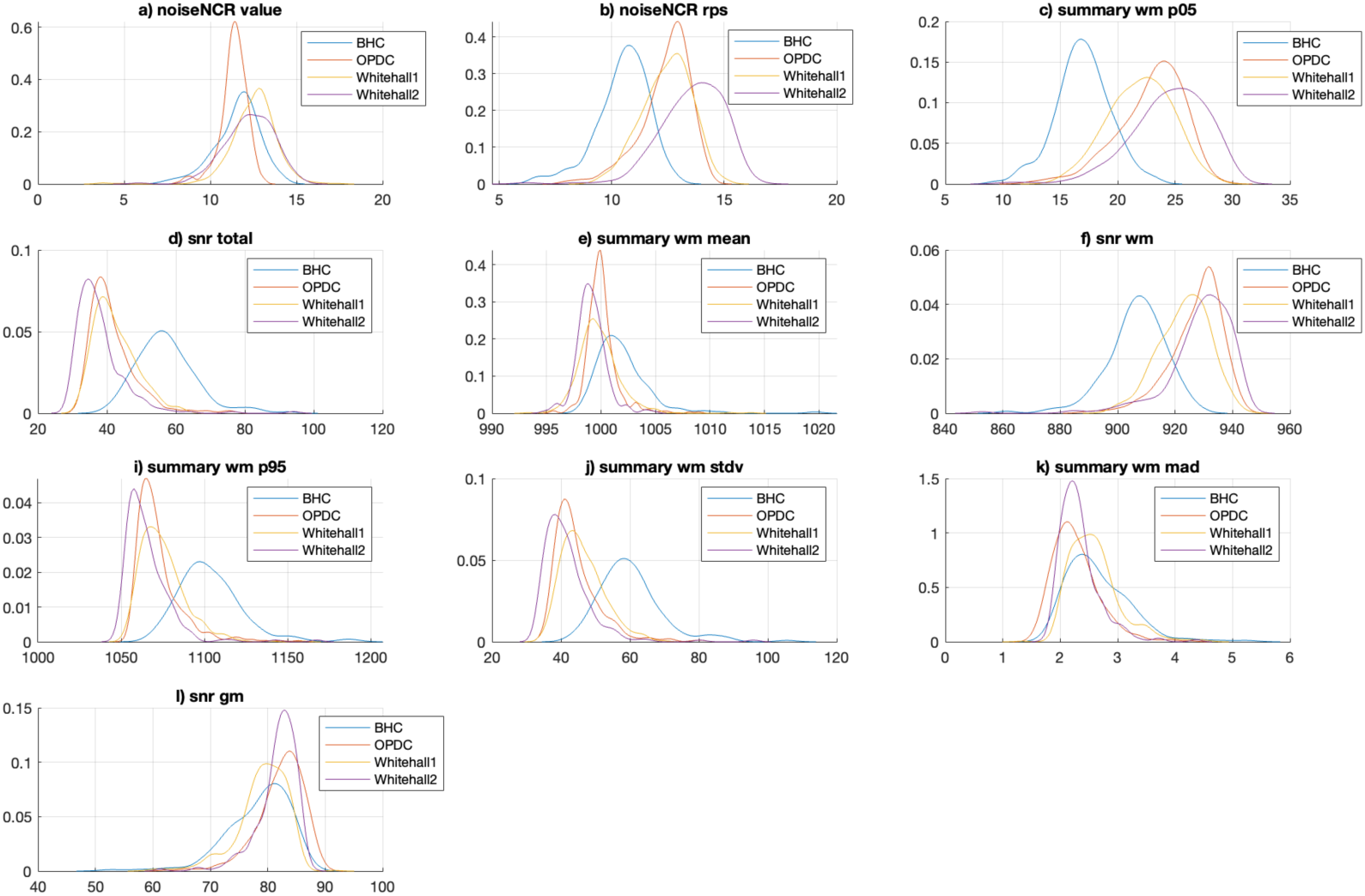
Kernel density plots showing the distribution of top 10 ranked features in final combined data model for BHC, OPDC, Whitehall1 and Whitehall2 sites. For a description of the features, please refer to tables Table 2 and Table 3.

The statistical significance test (KS-test) conducted on the 80 features showed notable differences in the distribution between the pairs of sites. Details of KS-test on each pair of sites are reported in the Supplementary material. Briefly, significantly different distributions were observed for various features (>40% of 80 features) between the ADNI sites (Siemens – 1.5T, 3T, GE – 1.5T, 3T, Philips – 1.5T, 3T) with the BHC, OPDC and Whitehall sites. When comparing the distribution within ADNI sites very few (<13% of 80 features) or none of the features showed significantly different distribution between the pairs of sites. Additionally, when comparing the distribution of features within non-ADNI sites (BHC, OPDC, Whitehall), many features (>67% of 80 features) showed statistically significant distribution.

As an example, **Figure 9** presents scatter plots illustrating the relationship of two features: noiseNCR_rps (CAT12) and snr_total (MRIQC). These plots offer insights into the distribution patterns of these features across various scenarios. Noticeable clustering is observed between two different datasets (BHC from Siemens 3T and ADNI GE 1.5T), acquired from distinct scanner manufacturers and field strengths. However, there is no clustering within the sites of ADNI dataset irrespective of the difference in the scanner manufacturer (ADNI Philips 3T and ADNI Siemens 3T) and field strength (ADNI Siemens 1.5T and ADNI Siemens 3T). Another notable observation is the evident clustering observed between the BHC and Whitehall1 datasets, despite both datasets being acquired using Siemens 3T Prisma scanners.

**Figure 9.**
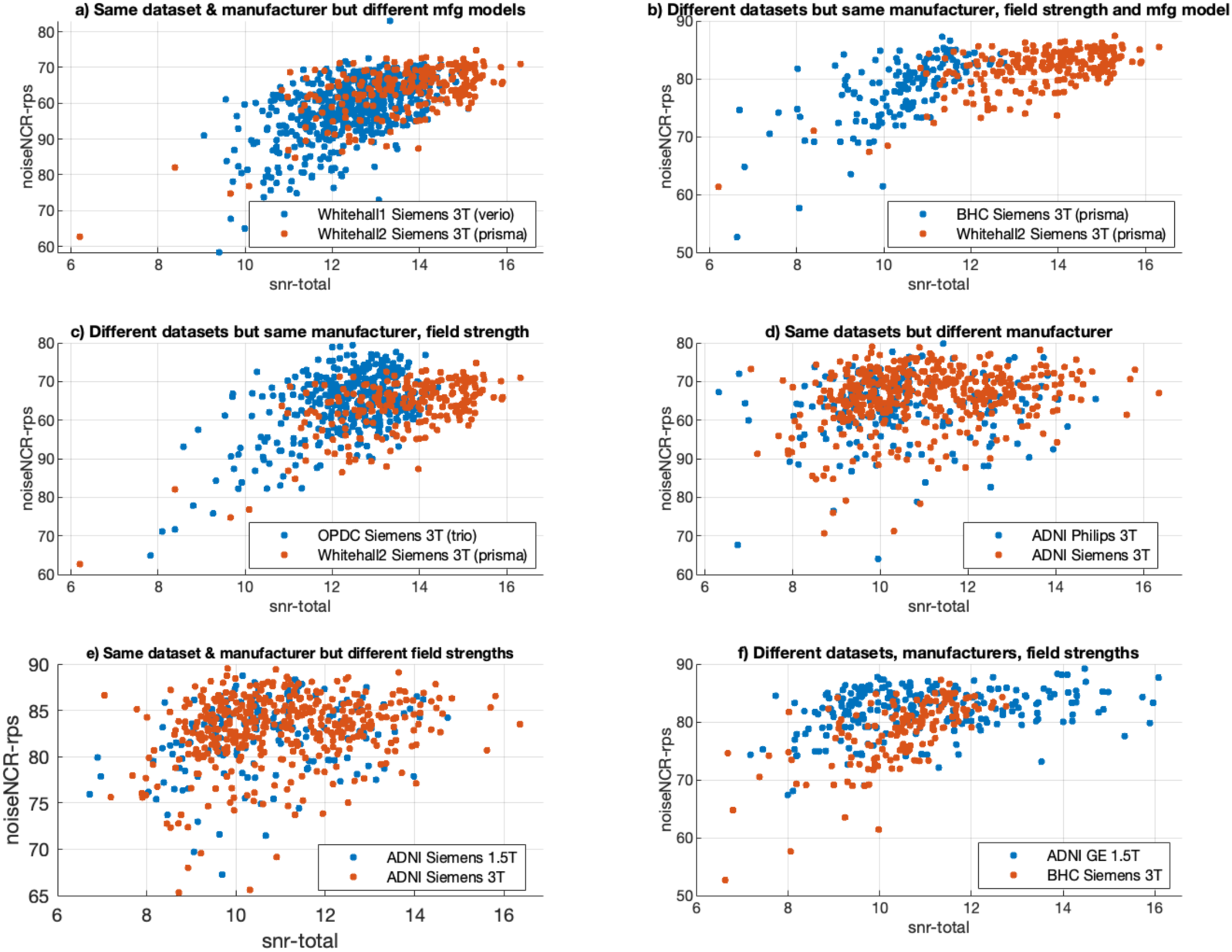
Scatter plots for two features snr-total from MRIQC on x-axis and noiseNCR-rps from CAT12 on y-axis showing different levels of overlap for different combinations of dataset, field strength and manufacturer.

### Leave-one-site out models

From the results on the combined model, the RUS classifier gave the best performance and was used for further experiments. Across all sites, the proposed RUS classifier achieved the highest balanced accuracy (78.2 ± 8.3 %) as compared to MRIQC (67.5 ± 11.5 %) and CAT12 (60 ± 7.2 %) (**Figure 10**). When comparing the balanced accuracy for each site, the proposed RUS classifier consistently performed better than MRIQC except for two sites (ADNI Philips 1.5T, OPDC Siemens 3T) where it showed 1% lower balanced accuracy than MRIQC. As expected, the balanced accuracy for individual sites in leave-one-site-out models tended to be lower compared to the results from the combined data model (average across sites = 85.6 ± 10 %, displayed for reference in Figure 10), due to fewer samples available in the test data and the presence of site-specific data in the training set for the combined data model.

**Figure 10.**
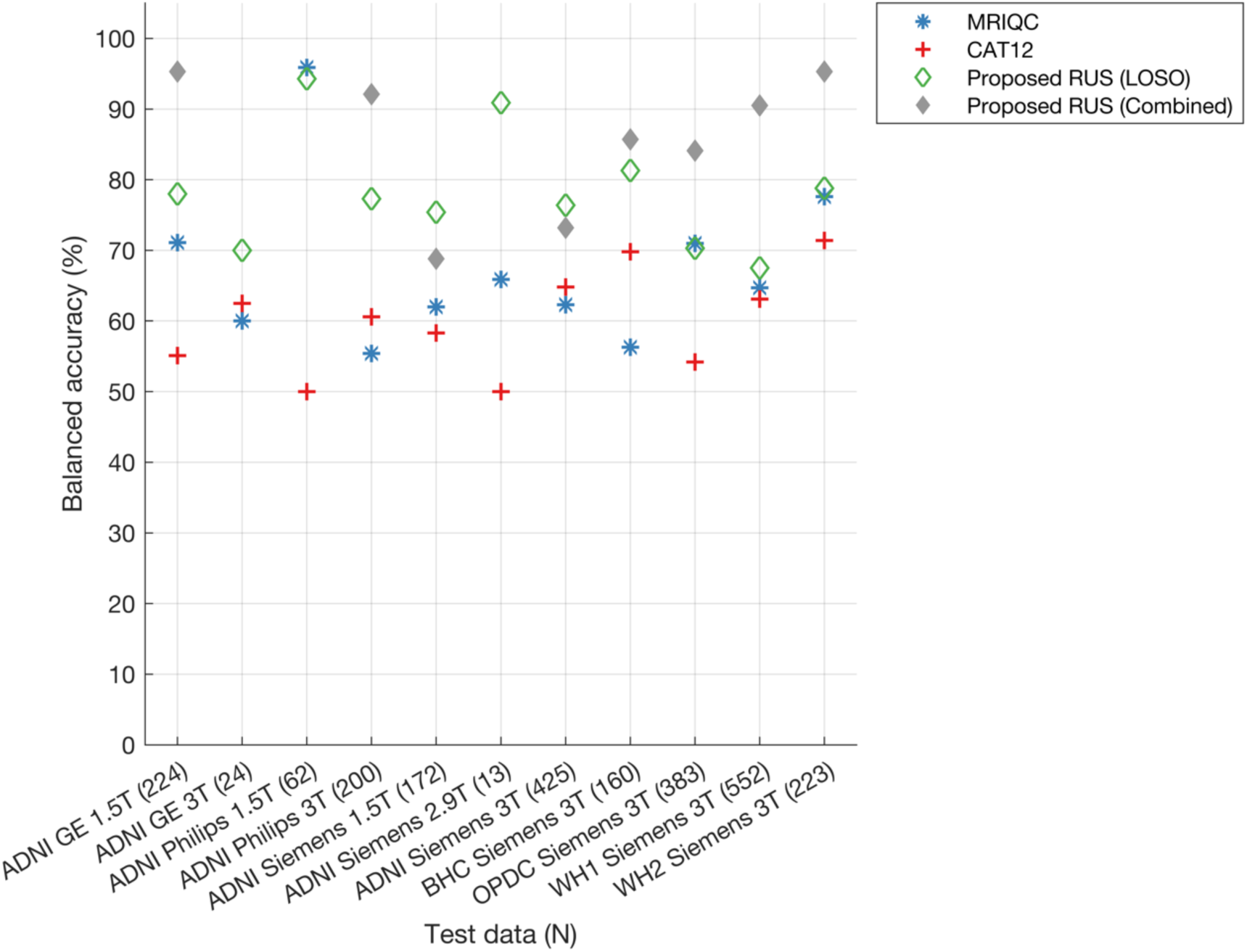
Balanced accuracy of MRIQC, CAT12 and the proposed RUS classifier. The total number of samples for each test site are provided in brackets (x-axis). For RUS classifier, each site was kept as test data and classifier was trained on remaining sites using the hyperparameters and feature ranking derived from combined data model (best model with 80 Feature size). For reference, we also provide the balanced accuracy of RUS classifier for each site within the test data of the combined data model to see how well our classifier generalises to test data from different sites (diamond marker with grey colour). Note that balanced accuracies for the combined data model are not included for three sites (ADNI GE 3T, ADNI Philips 1.5T and ADNI Siemens 2.9T) due to the absence of samples in the reject class of the test data (resulting in NaN values for balanced accuracies)

### Exploratory models

For all three exploratory models, the proposed RUS classifier consistently showed the highest balanced accuracies (73.8% - 80.4%) compared to MRIQC (63.8% - 67.9%) and CAT12 (56.6% - 58.3%) (**Figure 11**). Additionally, when comparing performance across exploratory models, the model trained on 3T scanners and tested on 1.5T scanners data showed higher balanced accuracy (80.4%) than the other two cases (manufacturer = 78.9%, field strength and manufacturer = 73.8%), probably due to the higher number of training samples (See **Table 7**). The ‘manufacturer’ model trained with Siemens data (1.5T, 2.9T, 3T) showed 84% balanced accuracy on Philips scanner data (1.5T, 3T) and 75% balanced accuracy on GE scanner data (1.5T, 3T). The model trained with 3T Siemens data (Field strength + Manufacturer) showed 72.4% balanced accuracy on test data from Siemens scanner (1.5T, 2.9T), 73.3% balanced accuracy on test data from GE scanner (1.5T, 3T) and 76.6% balanced accuracy on test data from Philips scanner (1.5T, 3T).

**Figure 11.**
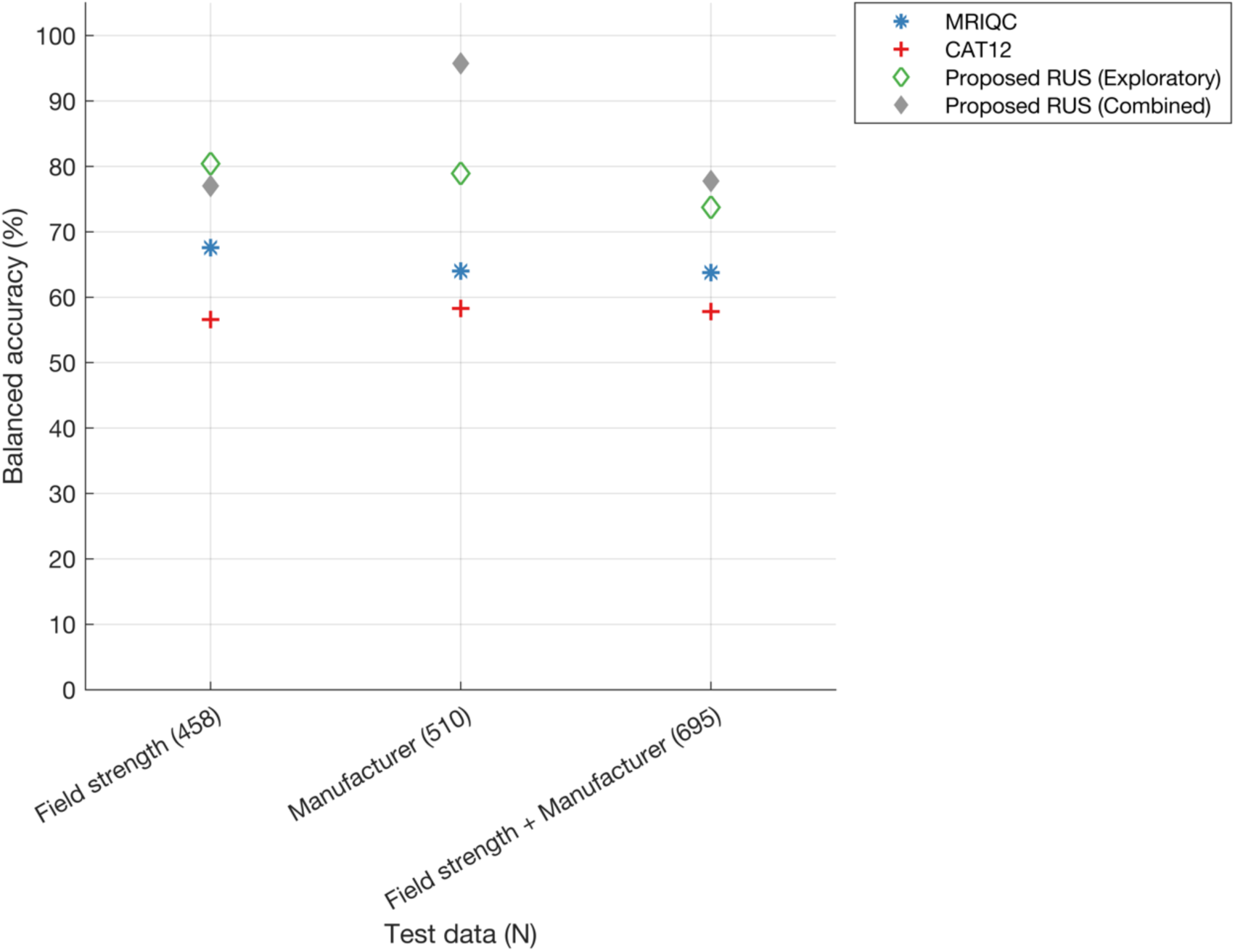
Balanced accuracy of MRIQC, CAT12 and the proposed RUS classifier for exploratory models. The total number of samples for each test site are provided in brackets (x-axis). Field strength: performance of models trained on 3T scanners data (Siemens, Philips, GE) and tested on 1.5T (Siemens, Philips, GE) and 2.9T (Siemens) scanners data; Manufacturer: performance of models trained on Siemens (1.5T, 2.9T, 3T) data and tested on Philips (1.5T, 3T) and GE (1.5T, 3T) data; Manufacturer and field strength: performance of models trained on Siemens 3T data and tested on Siemens (1.5T, 2.9T), Philips (1.5T, 3T) and GE (1.5T, 3T) data. Additionally, the balanced accuracy of the RUS classifier within the test data for the combined data model for each scenario is presented for reference (diamond marker with grey colour).

Also, in this case the performance on test data from the combined model for each of the three models (reported for reference in **Figure 11**) showed higher balanced accuracies (except field strength exploratory model which achieved 3.4% higher accuracy).

## Discussion

In this study we investigated approaches for automated quality control of T1w brain scans for ageing and clinical datasets acquired from multiple sites. The existing tools assessed in this study, MRIQC and CAT12, offer a broad array of quality metrics both from raw and processed images. We observed that some of the metrics are common between the tools, either assessing the same measures or highly correlated measures, while others are unique (i.e. not significantly correlated to measures from the other tool). When looking at the agreement in the accept or reject ratings between these tools and with visual QC, we found high variability across datasets, suggesting that these tools might not be suitable for highly heterogenous clinical datasets and the decision to accept or reject a scan will differ based on the dataset and the chosen tool. We observed enhanced agreement between visual QC and these tools after modifying the acceptance threshold. Nevertheless, these enhancements varied across different datasets, indicating that the adjusted thresholds may not be suitable for all clinical datasets. We then proposed a QC prediction approach by combining the quality measures from the automated tools to create a new classifier. The proposed RUS classifier exhibited higher performance than SVM and RF and good generalisability of prediction on the test datasets from diverse sites, scanner manufacturers and field strengths (balanced accuracy 87.7% on combined test data; average balanced accuracy 78 ± 8.3% on 11 test sites; average balanced accuracy 77.7 ± 3.5 % on exploratory models).

The RUS classifier outperformed MRIQC predictions and CAT12 QC ratings, supporting the benefit of using a combination of MRIQC and CAT12 quality measures. This is evident from the feature ranking, where the selected features at the top originated from both tools. Additionally, we explored the distribution of the quality measures that significantly contributed to the high performance (top ranked features) and observed that certain measures effectively captured variations across datasets (even for datasets acquired using scanners from the same field strength and manufacturer, for example, BHC and Whitehall2 datasets both acquired on Siemens 3T Prisma scanners). This highlights the complex technical differences among datasets, which might be influenced not solely by scanner manufacturer or field strength, but also by other factors for (e.g., acquisition parameters, number of channels in head coil, cohort characteristics such as age, sex, diagnosis etc.). These quality measures, when used in the context of harmonization techniques such as Neuroharmony (Garcia-Dias et al., 2020), could be instrumental in mitigating site-related effects in studies involving data from multiple sites.

A limitation of this study arises from the highly curated nature of the datasets, resulting in a significant imbalance between accept and reject labels. To address this, we focused on optimising balanced accuracy rather than overall accuracy. We also implemented multiple iterations of nested cross-validation (total 100) to iteratively validate and train our model on different samples. The use of the RUS classifier effectively addressed the issue of class imbalance by implementing under-sampling on the majority class (accept-labelled scans) to match it with the minority class (reject-labelled scans) during the training phase. This is clearly demonstrated by the improved specificity in predicting reject class labels, resulting in a notable enhancement in the balanced accuracy of RUS prediction when compared to RF, SVM, and other automated tools.

The RUS classifier achieved comparable performance on data from patients and controls (balanced accuracy of 86.8% and 88.3% respectively). We observed differences in performance across diagnostic subgroups, but while for ADNI the performance was lower in dementia than controls, in OPDC the performance was lower for controls than PD patients. This suggests that while diagnostic status of scans could affect results, the results may also be influenced by the total number of samples and number of scans in the reject class within each subgroup, making it difficult to perform a fair direct comparison across subgroups. We used a similar number of samples from both classes (accept and reject) in the training and test datasets for patients and controls, but not necessarily balanced within subgroups, due to the differences highlighted above. The performance across test sites (leave-one-site-out) and datasets in exploratory models indicates that our RUS classifier, when trained with different training datasets, maintains strong generalisation capabilities across diverse sites, scanners, and field strengths.

Another limitation of this study arises from the use of defaced T1w scans which involves the removal of facial features to protect individuals’ privacy. This step modifies the image, potentially altering the characteristics use for quality control. Recent studies have also indicated that defacing might influence the estimation of brain morphometry in contrast to non-defaced images (Bhalerao et al., 2022; Rubbert et al., 2022). While this issue remains an ongoing concern within the neuroimaging community, we decided to use defaced images as this is currently the best practice for sharing datasets and our goal was to develop a QC approach able to work on multiple datasets, likely aggregated from different sources on a data sharing and analysis platform, like the DPUK portal. For consistency, we applied the same defacing method (fsl_deface) across all datasets. Another constraint stems from the fact that the visual QC was performed by different raters, as we relied on visual QC ratings provided by the dataset owners. While this could have impacted the results as different raters may have had different subjective threshold for quality control, this setting reflects the real-world scenario of combining datasets from different sources. Nonetheless, our approach effectively captured the dataset variability and demonstrated high performance across all test cases, outperforming the other automated tools. Our primary goal was to develop a classifier using existing datasets available for sharing. However, as mentioned one of the challenges encountered is the limited availability of poor-quality scans (reject class), as shared datasets often are already highly curated. In future, obtaining more diverse and representative samples from the reject class could enhance the classification performance. Sharing poor-quality data can help the development of automated QC approaches, which can enhance the generalizability of classification on new datasets and ultimately to real-world clinical scans. Another strategy to address this issue involves leveraging synthetic image generation techniques. For instance, new datasets can be created by artificially introducing image artifacts into MRI scans derived from real-world data, thereby augmenting the sample size within the reject class (Ravi et al., 2024). However, the challenge is to create images that simulate realistic artefacts (Giuffrè & Shung, 2023). Another potential future direction would be to test the inclusion of more QC features (such as from FreeSurfer tool (Dale et al., 1999), UK biobank neuroimaging pipeline etc.(Alfaro-Almagro et al., 2018) in our classification framework to test if they result in increased performance (without significantly increasing the computational load) and/or further improve the generalisation of our classifier to new datasets. Our model is available to the community, and we plan to extend similar framework to test the quality of other MRI modalities.

## Conclusion

We proposed a classification model for quality assessment of T1-weighted scans of clinical datasets originating from diverse scanners, acquisition protocols, and spanning an elderly age range. Our approach involved combining the quality measures derived from automated tools, yielding promising performance, particularly when dealing with heterogeneous datasets from ageing and diseased cohorts. The code is readily available, and we will also share the QC metrics, trained classifiers, and outputs of this work through the DPUK data portal. This resource will serve as an asset for further exploration and robust QC of T1w scans across datasets, promoting comprehensive and reliable image quality assessment in future studies.

## Data Availability

Access to ADNI data is available to researchers upon request and approval of a data usage agreement (https://adni.loni.usc.edu/). Details on how to request access to the data can be found at http://adni.loni.usc.edu/data-samples/access-data/.
Other datasets used in this study can be accessed through the submission of an application via the DPUK data portal (https://portal.dementiasplatform.uk/Apply)

https://adni.loni.usc.edu/data-samples/access-data/

## Data Sharing

- Code is available here: https://git.fmrib.ox.ac.uk/mcz502/qc-paper
- Access to ADNI data is available to researchers upon request and approval of a data usage agreement (https://adni.loni.usc.edu/). Details on how to request access to the data can be found at http://adni.loni.usc.edu/data-samples/access-data/.
- Other datasets used in this study can be accessed through the submission of an application via the DPUK data portal (https://portal.dementiasplatform.uk/Apply)

## Acknowledgments

This work was supported by the UK Medical Research Council Dementias Platform UK (MR/T033371/1), an Alzheimer’s Association Grant (AARF-21-846366), the Wellcome Centre for Integrative Neuroimaging (203139/Z/16/Z), the NIHR Oxford Cognitive Health Clinical Research Facility and by the NIHR Oxford Health Biomedical Research Centre (NIHR203316). The views expressed are those of the author(s) and not necessarily those of the NIHR or the Department of Health and Social Care.

Work on the Whitehall II MRI substudy was funded by the Lifelong Health and Wellbeing Programme Grant: Predicting MRI abnormalities with longitudinal data of the Whitehall II Substudy (UK Medical Research Council: G1001354), the Horizon 2020 Grant: Lifebrain (Agreement number: 732592), and the HDH Wills 1965 Charitable Trust (No: 1117747).

Data collection and sharing for this project was funded by the Alzheimer’s Disease Neuroimaging Initiative (ADNI) (National Institutes of Health Grant U01 AG024904) and DOD ADNI (Department of Defense award number W81XWH-12-2-0012). ADNI is funded by the National Institute on Aging, the National Institute of Biomedical Imaging and Bioengineering, and through generous contributions from the following: AbbVie, Alzheimer’s Association; Alzheimer’s Drug Discovery Foundation; Araclon Biotech; BioClinica, Inc.; Biogen; Bristol-Myers Squibb Company; CereSpir, Inc.; Cogstate; Eisai Inc.; Elan Pharmaceuticals, Inc.; Eli Lilly and Company; EuroImmun; F. Hoffmann-La Roche Ltd and its affiliated company Genentech, Inc.; Fujirebio; GE Healthcare; IXICO Ltd.; Janssen Alzheimer Immunotherapy Research & Development, LLC.; Johnson & Johnson Pharmaceutical Research & Development LLC.; Lumosity; Lundbeck; Merck & Co., Inc.; Meso Scale Diagnostics, LLC.; NeuroRx Research; Neurotrack Technologies; Novartis Pharmaceuticals Corporation; Pfizer Inc.; Piramal Imaging; Servier; Takeda Pharmaceutical Company; and Transition Therapeutics. The Canadian Institutes of Health Research is providing funds to support ADNI clinical sites in Canada. Private sector contributions are facilitated by the Foundation for the National Institutes of Health (www.fnih.org). The grantee organization is the Northern California Institute for Research and Education, and the study is coordinated by the Alzheimer’s Therapeutic Research Institute at the University of Southern California. ADNI data are disseminated by the Laboratory for Neuro Imaging at the University of Southern California.

