## Supplementary materials 1 for "Automated quality control of T1-weighted brain MRI scans for clinical research: methods comparison and design of a quality prediction classifier"

### Performance of classifiers on combined test data

#### *Confusion matrix plots*

The confusion matrix for each classifier along with MRIQC and CAT12 is shown in **Figure S1**. The proposed RUS classifier showed the highest true negatives (reject labels correctly classified as reject) and the lowest false positives (reject labels wrongly classified as accept) as compared to all the other classifiers. The RF classifier showed the highest true positives (accept labels correctly classified as accept) and the lowest false negatives (accept labels wrongly classified as reject) as compared to all the other classifiers.

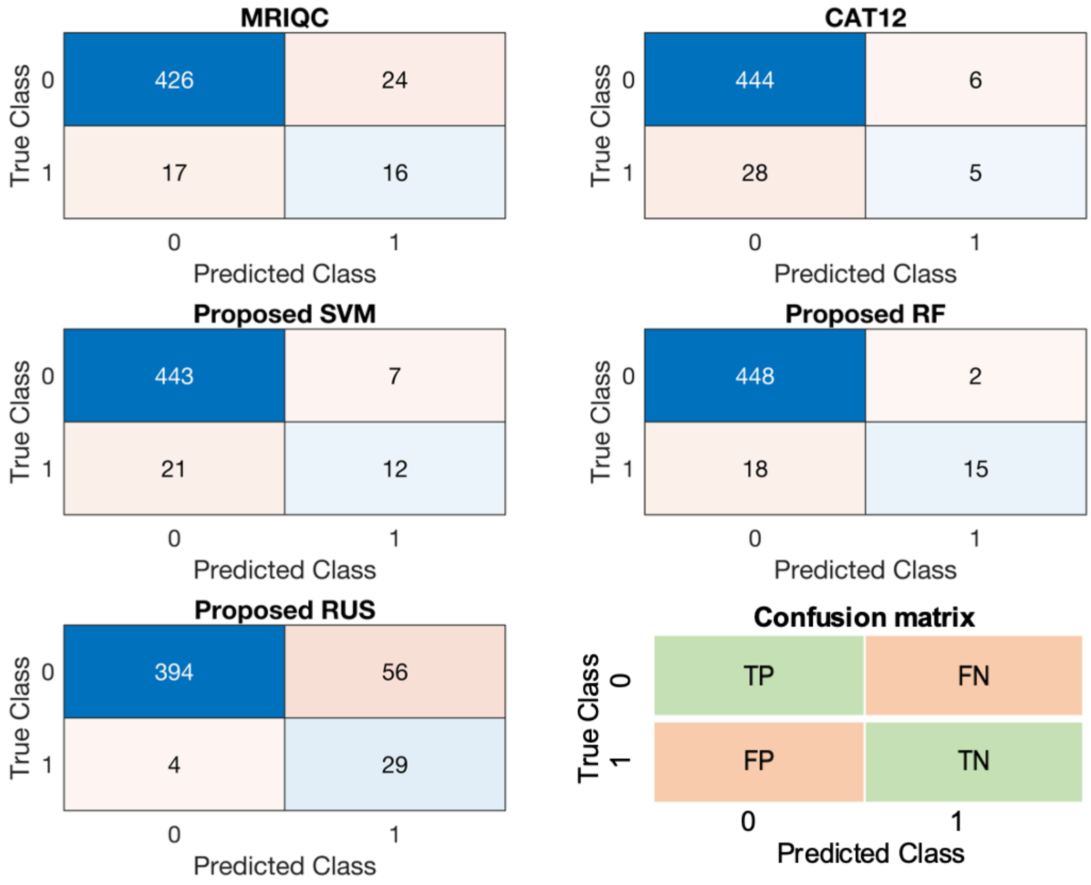


**Figure S1**. Confusion matrices showing total number of true positives, true negatives, false positives, and false negatives in the test data for MRIQC, CAT12 and proposed classifiers. Class 0 represents the accept class (positive), while class 1 represents the reject class (negative).

### *Performance across feature sizes*

The optimal feature size selected for SVM (balanced accuracy - 67.4%) and RF was 50 (balanced accuracy -72.5%), while for RUS was 80 (balanced accuracy - 87.7%)

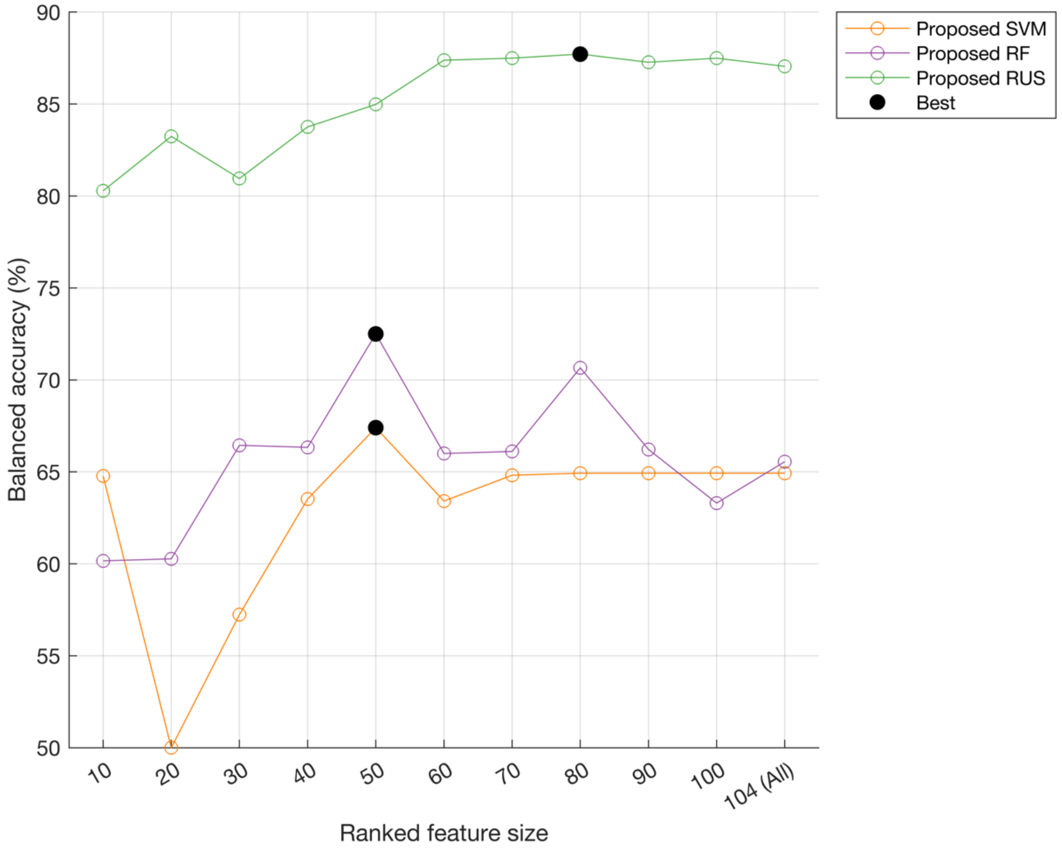


**Figure S2**. Balanced accuracy of proposed SVM, RF and RUS classifier on combined test data across different feature sizes.
